# Improving type 2 diabetes polygenic risk scores by incorporating rare, low-frequency, and population-specific variants

**DOI:** 10.1101/2025.11.24.25340878

**Authors:** Katie Taylor, Alicia Huerta-Chagoya, Xiaoyu Wang, Maheak Vora, Jiang Li, Lehte Turk, Teng Hiang Heng, Joohyun Kim, Andres Moreno-Estrada, Teresa Tusie-Luna, Carlos Aguilar-Salinas, Maggie Ng, Hilary C. Martin, David A van Heel, Genes & Health Research Team, Alisa Manning, David Carey, Priit Palta, Mitja I. Kurki, Sarah Finer, Tian Ge, Uyenlinh L. Mirshahi, Haoyu Zhang, Josep M Mercader

## Abstract

Polygenic risk scores (PRSs) can improve type 2 diabetes (T2D) risk prediction beyond clinical risk factors, but most exclude low-frequency, rare, and population-specific variants. We hypothesized that incorporation of rare variants via large-scale, well-imputed or whole-genome sequence-based genome-wide association (GWAS) meta-analyses and expanded linkage disequilibrium (LD) reference panels would improve risk prediction for T2D. We constructed a GWAS meta-analysis (230,675 T2D cases and 991,401 T2D controls), enabling the inclusion of rare variants (minor allele frequency [MAF] range= 1×10^−5^ - 0.01) to construct three T2D PRSs: (i) CTSLEB, which utilizes a custom ancestry-matched LD panel of 79.5 million variants and 83K participants to specifically model LD of rare variants; (ii) PRS-CS (TAGIT), using a reference panel expanded to 2.3 million variants to better capture low-frequency and population specific variants (population-specific MAF ≥ 0.01); (iii) PRS-CS (HM3), using a standard LD panel with HapMap3 variants (1.2 million variants). Performance was evaluated in the All of Us Research Program (20,301 T2D cases; 30,617 T2D controls) and compared to a benchmark multi-ancestry PRS (MAF≥0.01), developed by the D-PRISM consortium and derived from a significantly larger set of ancestry-specific meta-analyses (totaling 359,891 T2D cases and 1,825,792 controls). Expanding variant coverage with PRS-CS (TAGIT) and CTSLEB improved risk prediction relative to PRS-CS (HM3). While PRS-CS (TAGIT) showed greater prediction accuracy in the overall population, CTSLEB uniquely captured risk driven by rare variants, showing greater prediction accuracy for carriers of rare and low-frequency variants compared to PRS-CS (TAGIT) (AUC = 0.832 vs. 0.823 p(DeLong test) = 7.9×10^−5^) and PRS-CS (HM3) (AUC = 0.832 vs 0.818, p(DeLong test) = 2.39×10^−7^). The benchmark D-PRISM PRS showed the highest predictive performance for all ancestries except in African ancestry populations, where CTSLEB performed similarly for the overall population (CTSLEB AUC = 0.786 vs. D-PRISM AUC = 0.784, p(DeLong test) = 0.57) and significantly better for rare variant carriers (CSTLEB AUC = 0.775 vs. D-PRISM AUC = 0.768, p(DeLong test) = 8.71×10^−3^). These results demonstrate the value in incorporating rare and population-specific variants into PRS construction, improving genetic risk prediction in diverse populations.

## Introduction

Type 2 Diabetes (T2D) represents one of the current most significant global public health challenges. An estimated 10.5% of adults worldwide are currently diagnosed with T2D, with an additional 4.7% living with undiagnosed disease^1^. While environmental factors play major roles in T2D development, genetic susceptibility is also a key contributor^2–4^. Over a thousand genomic loci have been associated with T2D risk^5^, and over 100 polygenic risk scores (PRS) have been developed to identify individuals at increased risk^6^. PRSs estimate genomic risk for a complex trait or disease through the weighted sum of many variant effects across the genome, typically refining these effects by modeling linkage disequilibrium (LD) through clumping and thresholding^7^, penalization^8,9^, or Bayesian priors^10,11^.

Until recently, large-scale meta-analyses and PRS construction were restricted to common variants (minor allele frequency [MAF] > 0.005) due to limitations in genotype imputation and the availability of whole-genome sequence data. However, the heritability of T2D has notable contributions by rare and non-coding variants, with significant rare variant heritability enrichment outside of whole exome sequencing regions^12^. Recently, the largest rare-variant incorporating meta-analysis was developed, capitalizing on recent updates to TOPMed imputation. This study expanded the variant inclusion threshold to MAF ≥ 5×10^−5^ and identified eight rare (MAF < 0.001) independent signals significantly associated with T2D (p < 1×10^−8^)^13^. The effects of three high-impact variants located in genes known to cause monogenic diabetes of the young (MODY) were then evaluated in the context of common-variant PRS status. The impact of these variants differed by PRS status: carriers of an HNF4A variant in the highest PRS tertile showed an effect size comparable to that of carriers of a known MODY-causing variant in the same gene^13^.

Despite their importance, rare and low-frequency variants (MAF < 0.01) are historically excluded from PRS due to their omission from GWAS summary statistics and the sparsity of corresponding variants in LD reference used by PRS methods such as PRS-CS and PRS-CSx^11,14^. Recently, risk prediction performance has been increased via large scale rare variant association analyses and combination of monogenic gene risk and PRS scores^15,16^. Rare-variant integrated PRSs have also been developed by combining the effects of common-variant PRSs with aggregated rare-variant effects, typically derived through burden testing and ensemble learning^17,18^. This integration of rare variants into PRS shows clear improvements in predictive performance for continuous traits. However, these methods typically rely on access to individual-level whole-genome or exome sequencing data to define variant burdens. For binary traits like T2D, available sequencing datasets remain significantly smaller than the massive GWAS summary statistics currently accessible, which limits the power of these approaches to fully capture polygenic risk in broader populations^17^. Furthermore, while aggregating rare variants effectively boosts signal, it may occasionally mask the distinct risk profiles of individual high-impact variants.

We hypothesized that T2D risk prediction could be significantly improved by incorporating low-frequency and rare variants via large-scale GWAS meta-analyses and expanded reference panels. Accordingly, we conducted an expanded rare variant meta-analysis, comprising 230,675 cases and 991,401 controls from six ancestries and used the resulting summary statistics to develop PRS methods that allow incorporation of rare variants of large effects. We tested two complementary approaches: (1) **PRS-CS (TAGIT)**, utilizing PRS-CS with an expanded reference panel containing 2.3 million variants that improves coverage of low-frequency and population-specific variants^19^, and (2) **CTSLEB**, a method that is clumping-based, allowing it to utilize any custom LD reference panel to model local linkage disequilibrium. CTSLEB generates multiple variant weight sets using different LD thresholds, evaluates their predictive performance in a training population, and derives final variant weights by linearly combining effect sizes through an ensemble learning algorithm in the training population^20^. Crucially, for CTSLEB, LD was estimated using a custom reference panel of 79.5 million variants, and 83,818 participants with ancestry proportions matching that of the meta-analysis, enabling rare variant inclusion and accurate LD estimation.

We evaluated these scores in the AoU version 8 cohort and compared them against a “benchmark” multi-ancestry PRS recently developed by the D-PRISM consortium. Notably, the D-PRISM benchmark was derived from a significantly larger meta-analysis (~360,000 cases vs. our 230,675 cases). While larger sample sizes typically dictate PRS performance for common variants, we demonstrate that our methodological expansion to rare variants allows CTSLEB to perform indistinguishably from the larger benchmark in African and African American ancestry populations and significantly better for carriers of rare variants in African ancestry populations. These results highlight that precise modeling of rare variation can complement sample size to improve genetic risk prediction in diverse populations.

## Methods

The strategy of this study is shown in Figure 1. Briefly, association analyses for our meta-analysis were conducted with genetic data from United Kingdom Biobank (UKBB)^21^, Mass General Brigham Biobank (MGBB)^22^, Genetic Epidemiology Research on Adult Health and Aging (GERA)^23^, Genes and Health (G&H)^24^, FinnGen study (Finngen)^25^, Estonian Biobank (EstBB)^26^, MyCode Community Health Initiative (Geisinger)^27^, and version 7 of the All of Us Research Program (AoU)^28^. Participants in AoU portion of the meta-analysis were limited to the Version 7 release of data to avoid overlap between the meta-analysis cohort and final testing-cohort (Figure 1A). PRS training was conducted with genetic data from a Latino-based cohort including The Slim Initiative for Genomic Medicine in the Americas (SIGMA), Mexican Biobank (MXBB), and The Mexican Metabolic Syndrome (METS). Additional released samples in AoU v8 were used in the final validation set with no first and second grade relatedness to AoU v7. In total, the meta-analysis included 230,675 T2D cases and 991,401 T2D controls. The meta-analysis included 90.6% participants of predominantly European ancestry, 1.8% participants of predominantly African and African American ancestry, 1.6% participants of predominantly Admixed American ancestry, 0.5% participants of predominantly East Asian ancestry, 5.0% participants of predominantly South Asian ancestry, and 0.1% participants of Middle Eastern ancestry. We standardize population descriptors for ancestry in all analyses by using consistent acronyms: African/African American (AFR), Admixed American (AMR), East Asian (EAS), European (EUR), South Asian (SAS), and Middle Eastern (MID). Ancestry labels oversimplify the continuum of human genetic diversity but are necessary at this stage to apply ancestry-specific PRSs and assess PRS performance across ancestries. The manuscript describing the discovery results from this meta-analysis are being prepared as a separate manuscript.

**Fig. 1.**
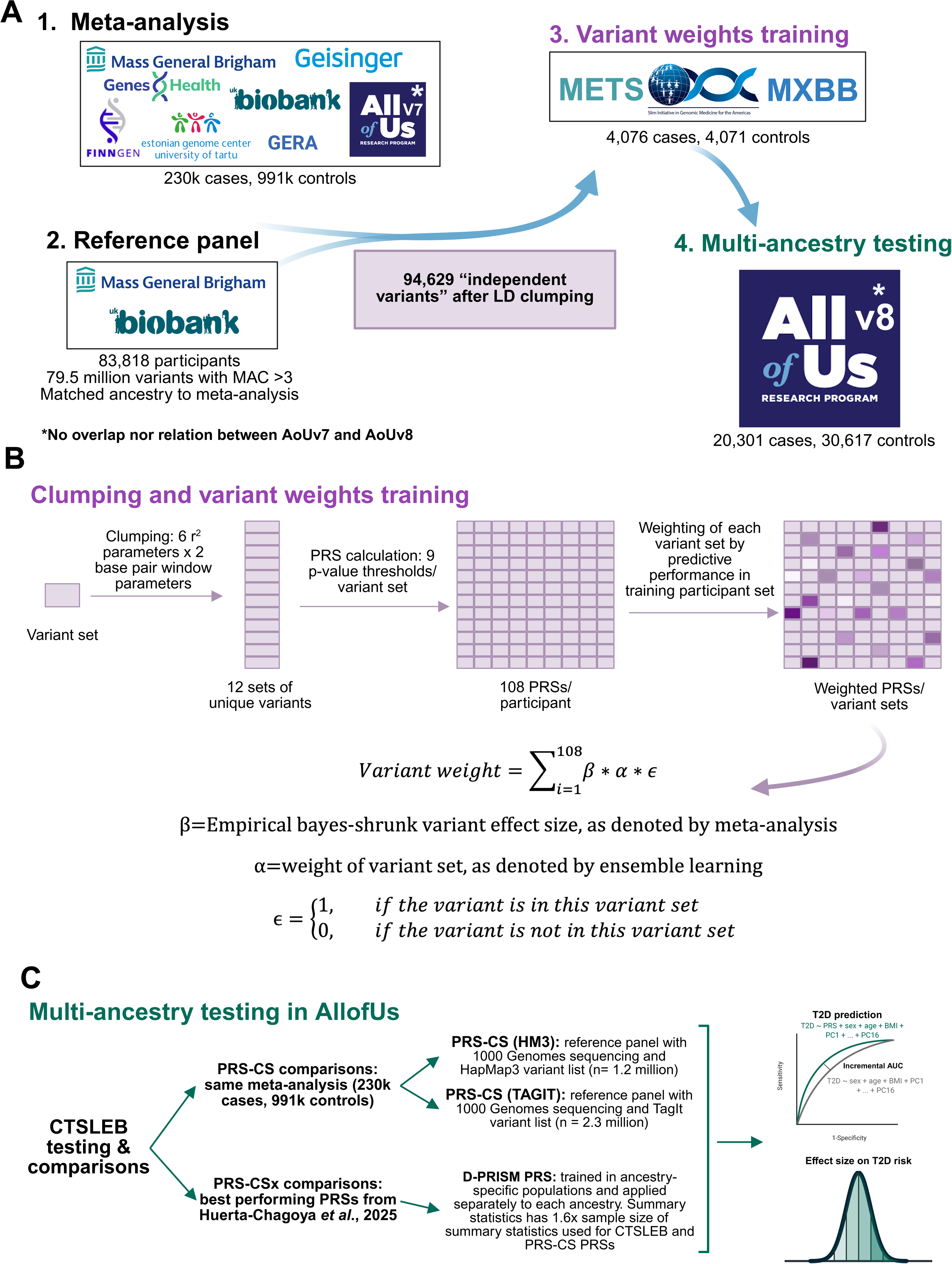
Overview of analysis workflow. A) *Meta-analysis and polygenic score construction.* We performed a meta-analysis of T2D using eight TOPMed-imputed or whole genome sequenced GWAS datasets (~230,000 cases and 991,000 controls). Independent T2D-associated variants were identified using an ancestry-matched LD reference panel generated from TOPMed-imputed data. Variant weights for the rare-variant–incorporating polygenic score (CTSLEB) were trained in a Latino cohort and evaluated in the AoU v8 dataset. B) *Clumping, thresholding, and empirical Bayes shrinkage.* We applied 12 clumping settings (6 LD thresholds, r2=0.01–0.8; × 2 window sizes, 62.5 kb–2000 kb) and 9 P-value thresholds (5×10^−11^ to 5×10^−4^), producing 108 clumped variant sets and 108 PRSs for the training dataset. Predictive performance of each score determined the weight of its variant set. Final CTSLEB variant weights are computed as a linear combination of the empirically shrunk effect sizes and the performance-based variant set weights. C) *Comparison of polygenic scores and performance evaluation.* CTSLEB was compared against two PRS-CS–derived scores generated from the same meta-analysis: PRS-CS (HM3), using HapMap3 variants (~1.2M), and PRS-CS (TAGIT), using the TagIt expanded variant panel (~2.3M). CTSLEB was also compared with “best-performing” D-PRISM polygenic scores generated from a meta-analysis with approximately twice the sample size. Predictive performance was assessed using: (i) incremental AUC, defined as the difference between AUC of the full model (PRS + age + BMI + principal components) vs. the covariate-only model (age + BMI + principal components), and (ii) odds ratio (OR) per standard deviation (SD) of the PRS distribution or OR comparing PRS percentile bins to the 40th–60th percentile reference group. Figure was created in https://BioRender.com.

### Cohort-specific information

UKBB is a prospective cohort study with DNA sequencing from ~500,000 individuals from across the United Kingdom between 2006 and 2010 with ages ranging from 40-69 years old. Participants gave information about their lifestyles, environment, and medical history at recruitment and agreed to have measurements taken and have health followed. The UKBB has obtained ethical approval covering the present study from the National Research Ethics Committee (REC ref. no. 11/NW/ 0382) and the data were accessed through application no. 27892. T2D cases (n=27,323) and controls (n=359,916) were defined using a UKBB-specific algorithm^29^.

MGBB, formerly Partners HealthCare Biobank, is a large data and biospecimen repository linked to electronic health records (EHR), containing DNA sequencing and linked measurements and EHR data from >65,000 patients from the MGB healthcare network. MGBB participants have provided consent for samples and data to be used in broad-based research. The approval for the analysis of MGBB data was obtained from the MGB Institutional Review Board (IRB; study no. 2016P001018). T2D cases (n=6,623) and controls (n=41,411) were defined with a curated algorithm utilizing structured and unstructured data^30^.

The GERA cohort was developed through the Kaiser Permanente Research Program on Genes, Environment, and Health (RPGEH) in partnership with the University of California, San Francisco (UCSF) Institute for Human Genetics (grant AG036607; Schaefer and Risch, principal investigators). This resource enabled GWAS in >100,000 adult participants enrolled in the Kaiser Permanente Medical Care Plan in Northern California who consented to participation in RPGEH. The Institutional Review Boards for Human Subjects Research of both Kaiser Permanente Medical Care Plan (Northern California Region) and the University of California at San Francisco approved the project, and data for the cohort were accessed through dbGaP (accession number phs000674.v1.p1). T2D cases (n=10,235) and controls (n=54,335) were defined by ICD-9-CM codes, as described on the dbGaP study page (https://www. <u>ncbi.nlm.nih.gov/projects/gap/cgi-bin/GetPdf.cgi?id=phd004308</u>).

The G&H study is a longitudinal, community-based study of British South Asian adults with >50,000 participants. Participants aged 16 years or older provided consent for broad-based research, completed questionnaires, consented to linking primary and secondary EHRs and national databases and consented to DNA sequencing. G&H was approved by the London South East NRES Committee of the Health Research Authority (14/LO/1240). T2D cases (n=10,631) and controls (n=44,186) were defined using EHR-derived clinical codes, following the study’s established phenotype algorithms.

The FinnGen study a national-scale biobank initiative in Finland integrating genomic data with longitudinal EHR data. This study contains DNA sequencing and linked measurements and EHR data from approximately 80,000 patients from across Finland. Genotype data have been generated on a custom array (~700,000 markers) followed by imputation using a Finland-specific reference panel to derive ~21 million variants per individual. Ethical clearance was granted by the Coordinating Ethics Committee of the Hospital District of Helsinki and Uusimaa (approval number HUS/990/2017) and by the Finnish Institute for Health and Welfare (permit numbers: THL/2031/6.02.00/2017; THL/1101/5.05.00/2017; THL/341/6.02.00/2018). T2D phenotyping for case (n=82,878) and control (n=403,489) phenotypes were defined as previously described in FinnGen publications^25^.

EstBB is a population-based cohort housed at the Institute of Genomics, University of Tartu, Estonia, and comprises >200,000 adult participants who provided broad informed consent and donated biospecimens and health-questionnaire information and agreed to link national health and insurance registers. Ethical approval for individual-level data analysis was granted by the Estonian Committee on Bioethics and Human Research (approval number 1.1-12/624). T2D case (n=14,704) and control (n=103,252) phenotypes were defined with ICD-10 and ATC coding systems.

The MyCode Community Health Initiative is a healthcare-system–based biobank launched by Geisinger, which enrolls patients throughout the Geisinger network, linking genomic data to EHRs. Participants consent to genotyping, broad research consent, and allow longitudinal follow-up of their health data. Informed consent, including permission for returning actionable genomic results and research use of data, is obtained from all participants. Ethical approval for MyCode and for related genomic research has been granted by the Geisinger Institutional Review Board (IRB numbers 2006-0258 and 2023-1149). T2D cases (n=52,010) and controls (n=41,639) were defined using EHR-derived criteria^13^.

The AoU research program is a large U.S.-based biobank from the National Institutes of Health, aimed to utilize the wide range of backgrounds in the US to facilitate and improve large genetic and epidemiological studies. We used the AoU short-read WGS data from cutoff date July 1, 2022 (release v.7) with short read whole genome sequencing for >245,000 participants for the meta-analysis. We used the AoU short-read WGS data from cutoff date October 1, 2023 (release v.8) with short read whole genome sequencing for ~447,000 participants for testing of PRSs. Phenotyping for T2D cases and controls for v.7 was determined with a modified version of the Northwestern T2D algorithm. Phenotyping for T2D cases (n=49,656) and controls (n=53,564) for v.8 was determined with a modified version of the algorithms utilized for MGBB^30^.

The SIGMA consortium is a Latin-America-focused genomic medicine initiative by the Carlos Slim Foundation and Broad Institute and collaborating Mexican and U.S. institutions, aiming to identify genetic risk factors for T2D and other conditions in Mexican and Latin American populations. More than 13,000 participants have consented to genotyping and broad research. Research has been approved by the Comité de Ética e Investigación del Centro de Estudios en Diabetes, the Ethics and Research Committees of the Instituto Nacional de Ciencias Médicas y Nutrición Salvador Zubirán, the Ethics and Research Committes of the Instituto Nacional de Medicina Genómica and the Federal Commission for the Protection against Health Risks (COFEPRIS) (CAS/OR/CMN/113300410D0027-0577/2012).

The MXBB project is a nationwide genomic resource designed to capture genetic diversity of the Mexican population and support biomedical research. Approximately 6,000 participants have consented to genotyping with linked health and trait data with written informed research consent. MXBB studies involving human participants were reviewed and approved by the Research Ethics Committee (Approval CI-1479) of the National Institute of Public Health, Mexico. The METS cohort is a prospective three-year observational cohort including Mexican adults living in large urban settings of central Mexico. Approximately 6,000 participants consented to genotyping and provided informed written consent for research. Research has been approved by the Ethics Committee of the Instituto Nacional de Ciencias Médicas y Nutrición.

Full information about each cohort is available in supplementary information (Fig 1a, Supplementary Tables 1,2).

For UKBB, MGBB, GERA, Geisinger, SIGMA, and MXBB, we separately applied quality control, phased, and imputed to the TOPMed r2 freeze 8^31^. Imputation was not necessary for AoU whole genome sequencing. Imputation was performed with TOPMed r3, Estonian Biobank-specific panel^32^, and Finngen-specific panels, for G&H, EstBB, and Finngen respectively. Variants were filtered to have r^2^≥0.3 for UKBB, MGBB, GERA, Geisinger, SIGMA, MXBB, and METS cohorts, and association testing was performed using REGENIE, with T2D as a binary outcome. Further quality control and cohort-specific covariates are listed in Supplementary Table 2, and the meta-analysis was conducted with METAL^33^.

### CTSLEB PRS development

The CTSLEB framework combines clumping and thresholding procedures with empirical bayes calibration and ensemble learning to optimize variant level weights for polygenic risk prediction^20^. The method requires three independent datasets: (1) GWAS summary statistics that provide the initial effect estimates, (2) a tuning dataset used to select hyper parameters, construct PRSs and train ensemble learning model that combines the scores (3) an external testing dataset used to evaluate final performance assessment. The tuning dataset comprised three cohorts: the SIGMA cohort, MXBB cohort, and METS cohort (Figure 1A). This Latino-based external dataset had 4,076 cases and 4,071 controls. Genotyping was imputed with TOPMed, as described above. Final evaluation was carried out in participants with whole genome sequencing newly released in version 8 of AoU^28^. Participants related to participants in AoUv7 were removed from the cohort, and the top inter-related participants of the cohort were removed with the R igraph package by removing participants related with PI_HAT value > 0.1875^34^. The resulting cohort had 20,301 cases and 30,617 controls (Supplementary Tables 1,2).

To construct the ancestry matched LD reference panel, we assembled a large custom genotype array with ancestry proportions matching our meta-analysis. Participants (n=83,818) were randomly selected from UKBB and MGBB cohorts. Ancestry-specific quality control was performed with QCTOOL -snp-stats^35^, and variants with ancestry-specific imputation quality less than 0.8 were removed from genotyping of participants matched to that ancestry. Cohorts were merged and variants with minor allele counts (MAC) below 3 were removed from the reference panel, resulting in 79,485,635 variants with MAC > 3 (Fig 1A).

The identification of unique variants and transformation of variant weights follows the CTSLEB PRS development framework^20^. After applying a p-value pre-filter threshold of p < 5×10^−4^, clumping and thresholding of variants identified in the meta-analysis was performed to select independent variants with 12 LD parameters. We applied a grid of R^2^ thresholds (0.01, 0.05, 0.1, 0.2, 0.5, 0.8) and base window sizes (50kb, 100kb). The clumping window was scaled inversely with the LD threshold 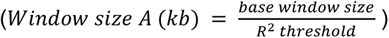 to ensure that lower R^2^ thresholds corresponded with larger genomic windows. This created 12 sets of unique variants. A combined matrix of effect size coefficients was created, assigning an effect size of 0 to variants in LD with other lead variants (Supplementary Figure 1). This effect size matrix was used alongside 9 p-value thresholds (5×10^−11^, 5×10^−10^, 5×10^−9^, 5×10^−8^, 5×10^−7^, 5×10^−6^, 5×10^−5^, 5×10^−4^) to assign 108 PRS scores per participant in the training cohort (Figure 1B).

After PRS scores for each variant set (n=108) were computed for each participant, linear models incorporating PRS scores and covariates (sex, age, BMI, 10 ancestry PCs) were created for each variant set. Variant weight sets were ranked based on predictive performance, measured by area under the curve (AUC), and variant sets with more than 0.98 correlation to the top performing variant set were excluded from further analysis. The R SuperLearner package with the SL.glmnet library was used to weight each variant set based on predictive performance^36^. Final variant weights were obtained by combining Empirical Bayes–calibrated effect sizes with variant set-specific weights (Figure 1B). Variants were given higher weights when they have higher individual association with T2D status, belong to more variant sets (are more independent), and belong to variant sets that produce PRSs more associated with T2D status. Final variant weights are distributable.

### PRS-CS PRS development

PRS-CS (HM3) and PRS-CS (TAGIT) were both developed with PRS-CS (version 1.1.0)^37^. PRS-CS infers variant effects by assuming a continuous shrinkage prior on variant effects from GWAS summary statistics. Both PRS-CS (HM3) and PRS-CS (TAGIT) utilized GWAS summary statistics from the same meta-analysis (230,675 T2D cases and 991,401 T2D controls) used to develop the CTSLEB PRS. Both methods used the software’s default settings. PRS-CS (HM3) used the most widely used developer-provided 1000 Genomes LD reference panels with common HapMap3 variants (n=1.1 million). Whereas, PRS-CS (TAGIT) used a newly D-PRISM developed LD panel with the 1000 Genomes genotyping and an expanded (n=2.3 million) set of SNPs using the Tag(ging) It(erative) of SNPs in multiple populations (TAGIT) program^38^, which had been previously shown to give better performance to PRSs, particularly in non-European populations^19^ (Figure 1C).

### Comparison against “benchmark” D-PRISM PRS

Ancestry-specific PRS-CSx weights were derived from the best performing PRS-CSx algorithms in each ancestry reported in Huerta-Chagoya *et al*., 2025^19^. This study utilized summary statistics from 125 selected GWAS in three large consortia: the Diabetes Meta-analysis of Trans-ethnic Association Studies (DIAMANTE)^39^, the Million Veteran Program (MVP)^40^, and FinnGen^25^ to compute large meta-analyses for AFR, AMR, EAS, EUR, and SAS populations. A total of 359,819 T2D cases and 1,825,729 controls were included in these summary statistics, (effective sample size is 128,695; 82,013; 400,683; 1,486,934; 87,223; for AFR, AMR, EAS, EUR, SAS, respectively). LD was modeled the best predictive panels^19^ and TAGIT SNP list (D-PRISM PRS), and PRS construction parameters for PRS-CSx were turned in a participant sample size of 10,992 cases and 31,792 controls^19^. As weights are ancestry-specific, performance for the D-PRISM PRS are reported for ancestry-specific populations (Figure 1C).

### T2D and age of T2D onset Phenotyping Definitions in AoU

T2D status was estimated with EHR and survey data in AoU, using a modified version of the Northwestern T2D EHR Case Definition. If participants did not have a T2D diagnosis in EHR, they needed to have been prescribed a non-insulin diabetes medication and have a record of abnormal glycemic labs to be considered cases. If patients had a T2D diagnosis and were not prescribed insulin, they either needed to have been prescribed non-insulin diabetes medications or have a record of abnormal glycemic labs to be considered T2D cases. If patients had a T2D diagnosis code and had been prescribed insulin, they either needed to have been prescribed a non-insulin medication after their insulin prescription or needed at least two T2D diagnoses recorded in EHR to be considered cases. Patients who self-reported Type 2 Diabetes in a survey were also considered T2D cases. Controls were participants over the age of 45 with at least one glucose measurement, no record of abnormal glucose measurements (Fasting glucose > 100 mg/dL, HbA2C > 5.7%, or random glucose > 200 mg/dL), no self-reported Type 1 diabetes or diagnosis of any related condition, and no prescription of insulin nor any other diabetes medications.

Age of diabetes onset was estimated with EHR and survey data. The earliest date of altered glycemia (HbA1C≥6.5% or random glucose ≥ 200 mg/dL in an outpatient setting), having an ICD code for T2D, or being prescribed a diabetes-specific medication in an outpatient setting (insulin or non-insulin) was extracted. When available, the estimated date of T2D onset was contrasted with self-reported date of diagnosis. If the estimated onset date fell outside of the self-reported diagnosis window, the evidence was considered insufficient to establish a reliable onset time-point.

### PRS Performance

We evaluated model performance in AoU v.8 by maximization of incremental AUC, defined as the difference in AUC for the full model incorporating PRS score, sex, age, BMI, and sixteen AoU-provided genetic principal components (PCs) and the covariate model incorporating sex, age, BMI, and genetic PCs. We also estimated the effect size of the PRSs as the odds ratio per standard deviation (OR/SD) unit of the PRS and report association with T2D status in generalized linear models with p-values. We calculated OR for individuals in the 0-2.5th, 0-5th, 0-10th, 10th-20th, 20th-30th, 30th-40th, 60th-70th, 70th-80th, 80th-90th, 90th-100th, 95th-100th, 97.5th-100th percentile groups compared to the 40-60th percentile group. We report results for the full AoU v.8 population (20,301 cases and 30,617 controls), as well as carriers of rare variants and samples enriched with carriers of rare variants. For variants in MODY genes, rare variants were defined as those having log-odds > 0.3 or < −0.3, MAF < 0.03, and p-values for T2D association in the meta-analysis > 1×10^−6^. For variants not in MODY genes, rare variants were defined as those having log-odds > 0.3 or < −0.3, MAF < 0.03, and p-values for T2D association in the meta-analysis > 1×10^−9^ (Supplementary Table 4).

### Sampling of populations enriched in carriers of rare variants

Samples enriched with carriers of rare variants are 4x the number of carriers or 2x the number of carriers for variants that have less than or more than 20 carriers, respectively. Non-carriers in these samples are randomly selected over 100 iterations, and incremental AUCs are reported as the mean and standard deviation of iAUCs for these 100 iterations. Results for both the full-population and the rare variant carriers are subset by ancestry across five ancestries: AFR, AMR, EAS, EUR, and SAS. Ancestries with sample sizes < 1000 or < 300 are excluded from results for the full population (SAS, MID) and rare-variant carriers (SAS, MID, EAS), respectively. Normalized PRS distributions for carriers and non-carriers of rare variants for each PRS method are reported, with differences in distribution tested by the Wilcoxon rank-sum test. Estimated mean OR for carriers of rare variants was calculated with the following equation: *estimated mean OR = e^mean(rare variant carrier PRS) × PRS effect size^*. PRS scores were also associated with age of T2D diagnosis in a model containing sex, age, BMI, and genetic PCs. Association significance and effect per standard deviation between normalized PRS and age of onset are reported.

## Results

### Expanded TAGIT reference panel and CTSLEB improve multi-ancestry T2D PRS prediction

To assess the benefit of expanding reference panels using PRS-CS and the more computationally efficient CTSLEB method, we first compared all approaches using the same meta-analysis summary statistics as input, which included 230,675 T2D cases and 991,401 controls. The study had 70.6% power to identify variants with minor allele frequency (MAF) ≥ 5×10^−4^, OR ≥ 1.5, at an alpha level of 5×10^−8^. This meta-analysis identified 1,378,041 variants with p < 0.0005, before clumping, which were further used to train and develop the CTSLEB PRS.

We first compared PRS performance in the full population without stratifying by ancestry (ALL n=50,918), comprising six ancestry groups (EUR, EAS, AMR, AFR, MID, and SAS). Odds ratios (ORs) were estimated across PRS percentile strata and per–standard deviation (SD) increase of the PRS, and predictive performance was assessed using incremental AUC (iAUC) (Figure 2).

**Fig. 2.**
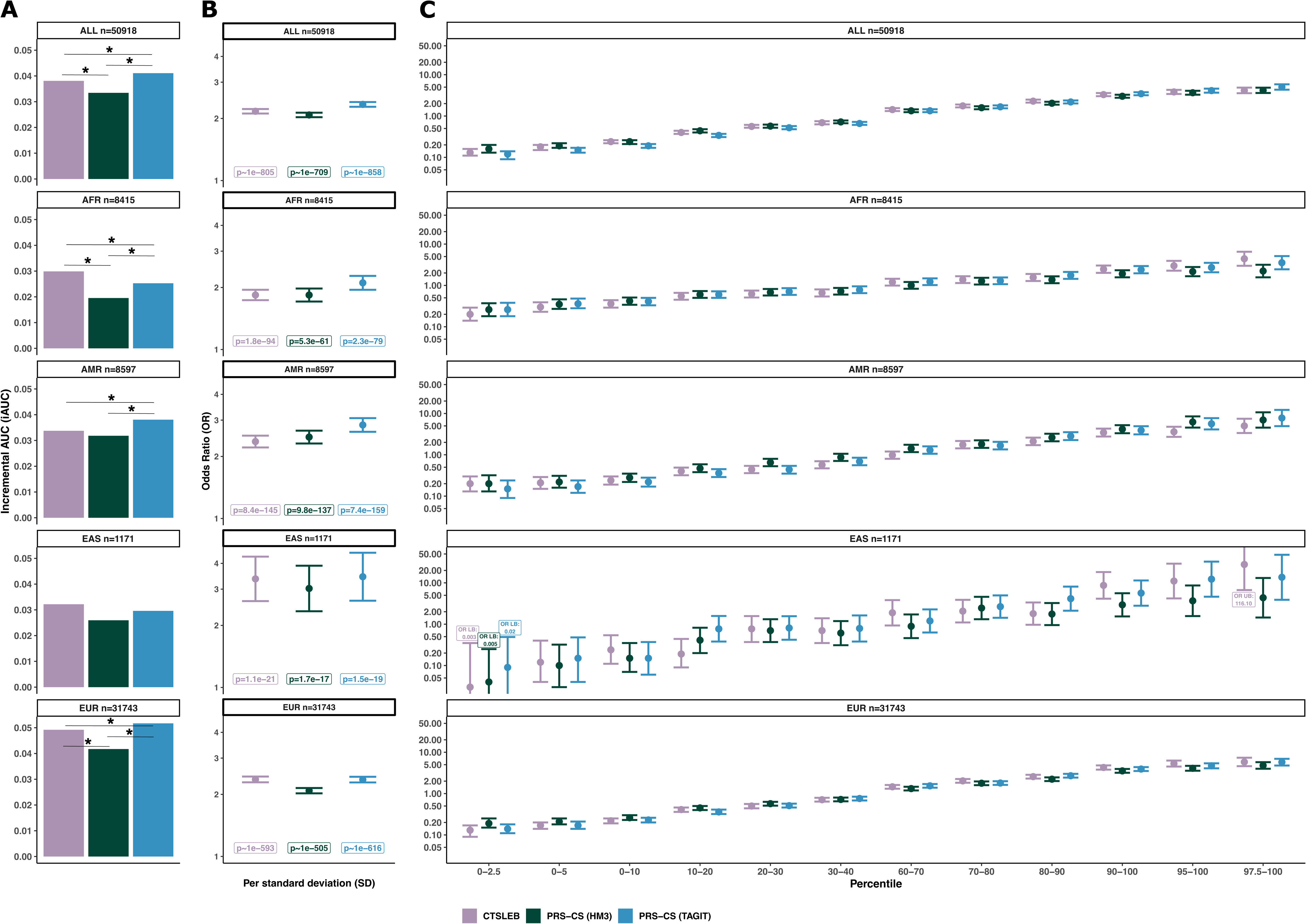
Performance of T2D PRSs utilizing the same meta-analysis in validation cohort for all participants and for five subset ancestry groups. **A)** Incremental AUC (iAUC) of T2D predictive performance for all participants and subset ancestries. Statistical significance in AUC difference tested with DeLong test, with significance (p **≤** 0.05) denoted by *. **B)** Odds ratio (OR) of T2D risk increase per standard deviation (SD) of PRS for all participants and subset ancestries. Association p-values written below each OR/SD estimate. **C)** OR comparing PRS percentile bins to 40th-60th percentile reference group for all participants, AFR, AMR, EAS, and EUR-subset populations. The Y axis is in log scale. Colors for all plots represent PRS used: purple for CTSLEB, green for PRS-CS (HM3), blue for PRS-CS (TAGIT). Error bars show 95% confidence intervals. Number of participants in each subset given above each plot.

Expanding the variant reference panel significantly improved risk prediction for PRS-CS. For the full multi-ancestry population, expanding the SNP list from PRS-CS from HapMap3 (1.2 million variants) to TagIt (2.3 million variants) improved performance (PRS-CS (TAGIT) AUC=0.832, iAUC=0.041 vs PRS-CS (HM3) AUC=0.817, iAUC=0.033, p(deLong test)=4.90×10^−45^) (Figure 2A, Supplementary Table 6). PRS-CS (TAGIT) was also more strongly associated with T2D than PRS-CS (HM3) [PRS-CS (TAGIT) OR/SD=2.34 (2.28-2.40), p~1×10^−858^ vs PRS-CS (HM3) OR/SD=2.08 (2.03-2.13), p~1×10^−709^] (Figure 2B, Supplementary Table 6). Furthermore, the highest PRS percentile groups for PRS-CS (TAGIT) had higher OR for T2D than the highest percentile groups for PRS-CS (HM3) (Figure 2C, Supplementary Table 6).

We next compared PRS performance in ancestry-subset participant populations. The small sample sizes of the MID (n=192) and SAS (n=584) cohorts, combined with their minimal contribution to the meta-analysis (0.4% and 0.8%, respectively), limited the ability to reliably evaluate PRS performance and were therefore removed from ancestry stratified analyses. In ancestry-stratified analyses, PRS-CS (TAGIT) outperformed PRS-CS (HM3) across all ancestry groups, with significantly larger AUCs for AFR, AMR, and EUR (AFR p(deLong test)=5.35×10⁻⁵, AMR p(deLong test)=4.90×10⁻⁷, and EUR p(deLong test)=3.54×10⁻⁴⁰).

Expanding the variant reference panel to incorporate rare variants with CTSLEB architecture significantly improved risk prediction relative to PRS-CS (HM3) with a standard reference panel size (CTSLEB AUC=0.830, iAUC=0.038 vs PRS-CS (HM3)=0.825, iAUC=0.033, p(deLong test)=8.20×10^−8^) (Figure 2A, Supplementary Table 6). Association strength measured by OR/SD (95% CI) followed the same pattern. CTSLEB was more strongly associated with T2D status than PRS-CS (HM3) [CTSLEB OR/SD=2.17 (2.03-2.22), p~1×10^−804^ vs PRS-CS (HM3) OR/SD=2.08 (2.03-2.13), p~1×10^−709^] (Figure 2B, Supplementary Table 6). The highest percentile groups for CTSLEB had higher OR for T2D status than the highest percentile groups for PRS-CS (HM3). CTSLEB also outperformed PRS-CS (HM3) in ancestry-stratified analyses, with significantly larger AUCs in AFR and EUR (AFR p(deLong test)=1.94×10⁻⁴ and EUR p(deLong test)=5.35×10⁻⁸).

CTSLEB showed the strongest association with T2D status in AFR (p=1.8×10⁻⁹⁴) and EAS (p=1.1×10^−21^), across the three methods, driven by more extreme ORs at the highest and lowest PRS percentiles. In contrast, PRS-CS (TAGIT) produced the highest OR per standard deviation and the most significant associations with T2D status in AMR (OR/SD=2.84; p=7.4×10⁻¹⁵⁹) and EUR (OR/SD=2.34; p~1×10⁻^616^). Notably, within EUR, individuals in the highest and lowest CTSLEB percentiles exhibited the largest and lowest ORs for T2D, respectively (Figure 2B,C; Supplementary Table 6).

### CTSLEB enhanced performance in modeling and predicting T2D risk in rare and low-frequency variant carriers

Following our hypothesis that including rare and low-frequency variants in PRS development would particularly improve modeling and performance in rare variant carriers, we subset the full population of AoU v.8 participants to only include rare variant carriers. We selected a subset of rare and low-frequency variants with large effect, defined as those with MAF < 0.03, log-odds effect size > 0.3 or < −0.3, and p < 1×10^−6^ or p < 1×10^−9^ for rare coding variants in monogenic diabetes genes or rare variants not monogenic diabetes genes, respectively. The selection of these variants was based on previous T2D association findings (Supplementary Table 4). Of the 3,773 participants who carried at least one of the selected rare variants, 2,036 carried risk increasing variants, 1,784 carried protective variants, and 47 carried both risk and protective variants.

#### Incorporating rare variants with CTSLEB allows the capture of risk change associated with carrying rare variants

We first compared PRS distributions between carriers and non-carriers of rare risk and protective alleles for each PRS method. CTSLEB showed stronger separation between carriers and non-carriers than either PRS-CS (TAGIT) or PRS-CS (HM3). For CTSLEB, PRS values were higher in carriers of rare risk variants than in non-carriers (Wilcoxon rank-sum p=1.50×10^−96^), corresponding to an estimated average OR based on the PRS value of OR = 1.44. CTSLEB assigned lower PRSs in carriers of protective variants, compared to non-carriers (Wilcoxon rank-sum p=6.29×10^−187^) with a mean estimated OR=0.57. In contrast, PRS-CS (TAGIT) and PRS-CS (HM3) showed no appreciable differences in PRS distributions between carriers and non-carriers, with mean estimated ORs in carriers ranging between 0.9 and 1.1 for both models (Figure 3A).

**Fig. 3.**
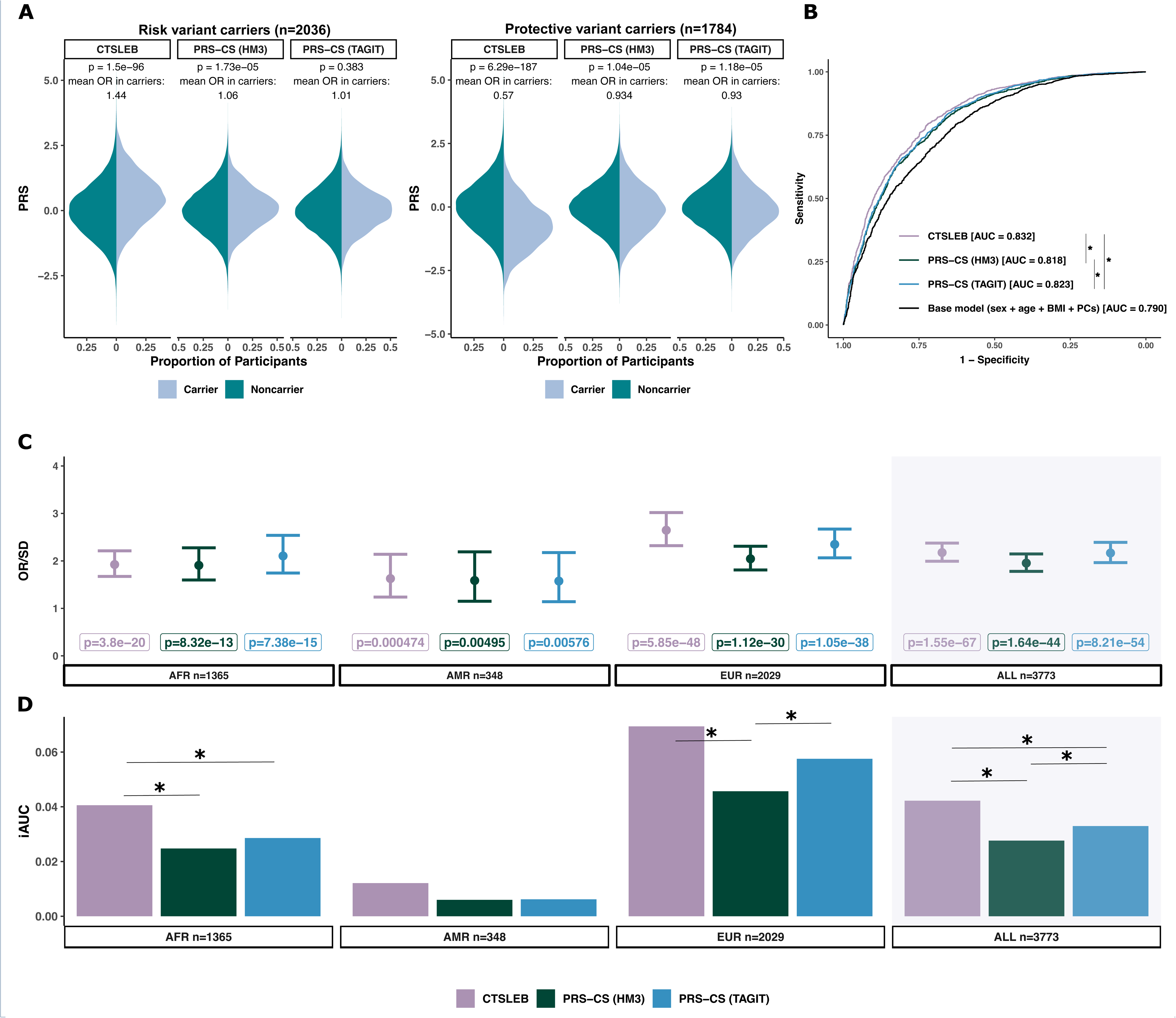
Performance of T2D PRSs utilizing the same meta-analysis in validation cohort for rare variant carriers. **A)** Mirrored PRS distributions for carriers versus non-carriers of risk and protective variants, respectively, for all three PRS methods. Significance in distribution differences between carriers and non-carriers tested by the Wilcoxon rank sum. P-values of these tests are written above each mirrored distribution. The mean PRS-estimated OR associated with being a carrier versus non carrier is also written above each mirrored distribution. Colors represent carrier status: dark teal for noncarriers, light blue for carriers. **B)** ROC-AUC curve of PRS models utilizing the same meta-analysis. AUC values are written in figure legend beside respective PRS models. **C)** Odds ratio (OR) of T2D risk increase per standard deviation (SD) of PRS for rare variant carriers for AFR, AMR, EUR sub-populations and all ancestries combined. Controls are rare variant carriers and T2D controls. **D)** Incremental AUC (iAUC) of T2D predictive performance for rare variant carriers in subset ancestries (AFR, AMR, EUR) and all rare variant carriers. Statistical significance in AUC difference tested with DeLong test, with significance (p **≤** 0.05) denoted by *. Colors for all plots represent PRS used: purple for CTSLEB, green for PRS-CS (HM3), blue for PRS-CS (TAGIT). Error bars show 95% confidence intervals. Number of participants in each subset given below each plot for C and D.

#### Incorporating rare variants with CTSLEB improves risk prediction in rare variant carriers

Given that CTSLEB captured more accurately the effects of rare variant carriers, we evaluated whether this translated to improved predictive performance among rare variant carriers. CTSLEB had higher predictive performance for T2D status in rare-variant carriers across the three methods (CTSLEB AUC=0.832, iAUC=0.042 vs PRS-CS (TAGIT) AUC=0.823, iAUC=0.032, p(deLong test)=7.9×10^−5^; CTSLEB AUC=0.832, iAUC=0.042 vs PRS-CS (HM3) AUC=0.818, iAUC= 0.028, p(deLong test)=2.39×10^−7^) in carriers of rare variants. CTSLEB also showed the strongest association with T2D status among rare variant carriers (OR/SD=2.18 (1.99-2.38), p=1.55×10^−67^) (Figure 3C, Supplementary Table 6). Expansion of the SNP list via PRS-CS (TAGIT) had higher predictive performance than PRS-CS (HM3) within this group (p=0.0191) (Figure 3B,D, Supplementary Table 6) and stronger association with T2D status.

We next stratified rare variant carriers by ancestry, reporting results for ancestry groups with > 300 rare variant carriers in the testing population (AFR, AMR, EUR). CTSLEB demonstrated the highest predictive performance in all three ancestries, with significant improvements in AUC against both PRS-CS models in AFR and against PRS-CS (HM3) in EUR (Figure 3D, Supplementary Table 6). CTSLEB also showed the most significant association with T2D status for AFR [OR/SD=1.92 (1.67-2.21), p=3.8×10^−20^] AMR [OR/SD=1.63 (1.24-2.14), p=4.7×10^−4^] and EUR [OR/SD=2.64 (2.32-3.02), 5.9×10^−48^] (Figure 3C, Supplementary Table 6).

CTSLEB also modeled increased risk carriers of individual rare variants and improved risk prediction accordingly for carriers of individual rare variants. Analysis of individual rare variant carriers was limited to variants with more than ten carriers in the validation cohort. We modeled PRS distribution for carriers of each individual variant versus all participants in all PRS algorithms and found that CTSLEB modeled the effect of all variants to a greater degree than either PRS-CS (TAGIT) or PRS-CS (HM3) (Figure 4A,B, Supplementary Table 7). CTSLEB PRS showed significantly higher PRS distributions in carriers of risk increasing variants and significantly lower PRS distributions in carriers of protective variants. For example, carriers of the p.Arg114Trp *HNF4A* variant, previously reported to be associated with 8-fold increased risk for T2D^13^ showed a significantly higher PRS compared to carriers (Wilcoxon rank-sum test p<1×10^−6^), with a mean estimated OR based on the PRS value of 3.1, with values ranging from 1.24 to 7.96 when using CTSLEB (Supplementary Table 7). However, the mean estimated OR remained closer to 1 when using PRS-CS (HM3) or PRS-CS (TAGIT). Both PRS-CS methods did not model the effect of rare variants in carriers, with mean estimated ORs for carriers (based on their PRS value) of all risk allele carriers remaining below 1.1 and mean OR for carriers of all protective alleles remaining above 0.91 (Figure 4B, Supplementary Table 7).

**Fig. 4.**
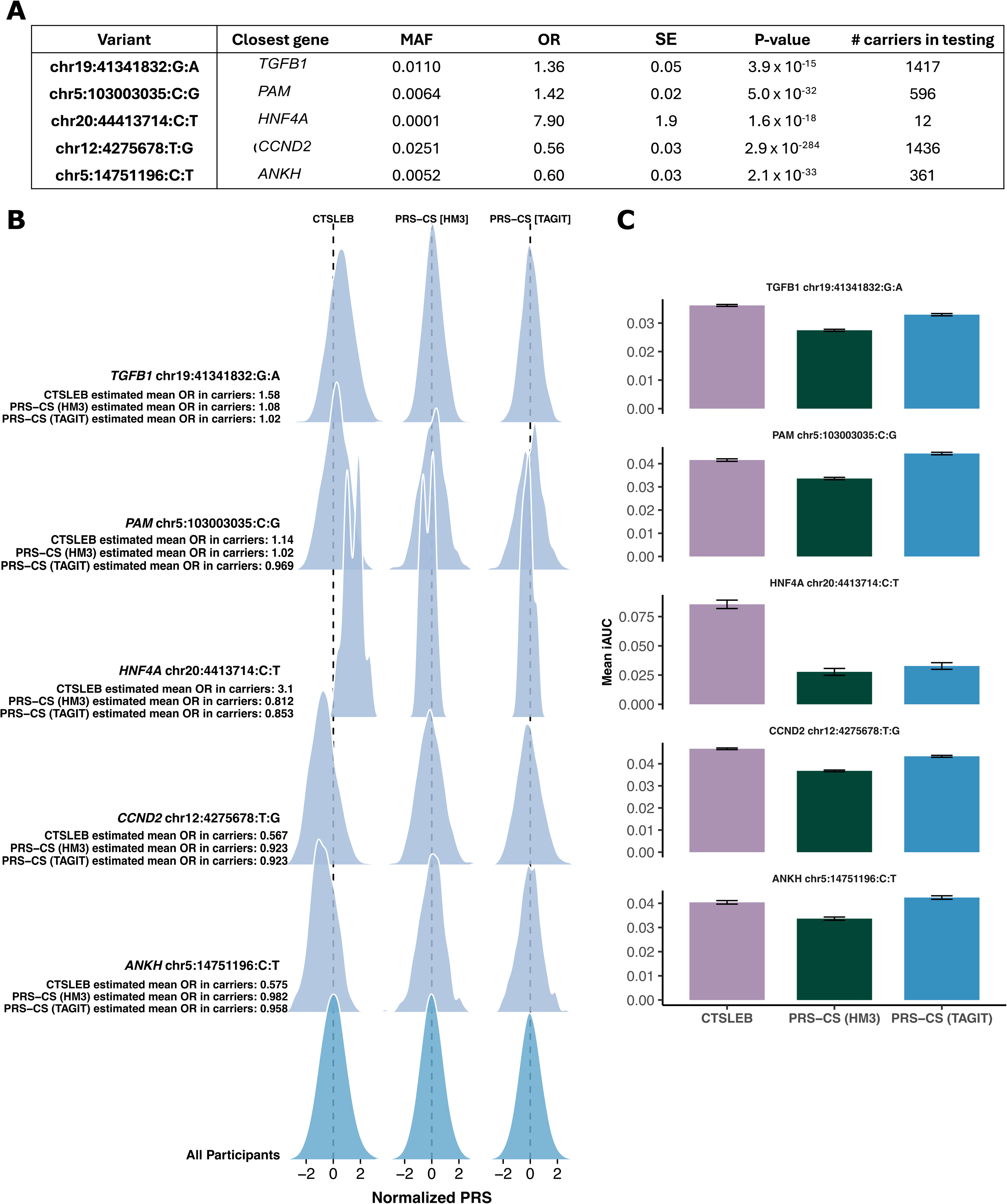
PRS distribution for carriers of individual rare variants and predictive performance for samples enriched with carriers of rare variants. A) Variants and associated MAFs, ORs, SEs, p-values, found by meta-analysis. Number of carriers in validation cohort. Position of variants is in b38. B) Ridgeline plot showing PRS distributions for carriers of rare variants for CTSLEB and PRS-CS comparisons created with the same meta-analysis. Variant and estimated OR for carriers by each PRS written to the left of distributions. Non-carrier distributions are at the bottom of the plot. Color represents carrier status: light blue for rare variant carriers and teal for non-carriers. C) Mean incremental AUC (iAUC) with 95% confidence interval for samples enriched with carriers. For variants with < 30 carriers in validation cohort, sample size is 4 x number of carriers. Otherwise, sample size is 2x number of carriers. Non-carriers are randomly selected and predictive performance for each sample set is tested over 100 iterations for each variant. Colors represent PRS used: purple for CTSLEB, green for PRS-CS (HM3), blue for PRS-CS (TAGIT).

To evaluate how modeling each variant influences predictive performance, we generated datasets enriched for rare variant carriers by selecting the carriers of interest and a random sample of non-carriers. For variants with fewer than 20 carriers, we randomly selected a comparison sample equal to three times the number of carriers. For variants with 20 or more carriers, we sampled a comparison group equal in size to the number of carriers. For each variant, this sampling procedure was repeated 100 times. Predictive performance is reported as the mean and standard deviation of iAUC values across all permutations. We found that CTSLEB performed best for samples enriched with carriers of variants in *TGFB1*, *HNF4A*, and *CCND2*. CTSLEB performed similarly well to PRS-CS (TAGIT) for samples enriched with carriers of variants in *PAM* and *ANKH.* CTSLEB performed better than PRS-CS (HM3) in all samples enriched for all rare variant carriers (Figure 4C, Supplementary Table 7). When subset by ancestry, CTSLEB had the highest mean incremental AUCs for AFR samples enriched with all individual rare variant carriers. PRS-CS (TAGIT) had the highest mean incremental AUCs for AMR and EUR samples enriched with carriers of all individual rare variants (Supplementary Table 7, Supplementary Figures 2-4).

### Benchmark D-PRISM PRSs with larger GWAS sample size outperform CTSLEB in full ancestry subsets

After comparing several PRS approaches developed with the same GWAS meta-analysis, and therefore, equivalent GWAS summary statistics sample size, we compared CTSLEB and PRS-CS (TAGIT) with the recently developed multi-ancestry PRSs by the D-PRISM consortium reported by Huerta-Chagoya et al. (2025)^19^, which were the top performing PRSs for all major ancestries (D-PRISM PRS). This study utilized summary statistics from 125 selected GWAS from three large consortia to compute large meta-analyses for AFR, AMR, EAS, EUR, and SAS populations. A total of 359,891 T2D cases and 1,825,792 controls were included in these summary statistics −1.6-fold increased sample size compared to the summary statistics used for previous methods- and the TAGIT SNP list (2.3 million variants) was used in LD modeling with ancestry-specific reference panels for each ancestry. PRS construction parameters for PRS-CSx were tuned in ancestry-specific populations in a total of 10,992 cases and 31,792 controls.

We compared PRS performance across four ancestry groups (EUR, EAS, AMR, and AFR). Predictive performance was measured with iAUC. Association of PRSs with T2D status was measured by OR/SD and association significance (Figure 5A-C).

**Fig. 5.**
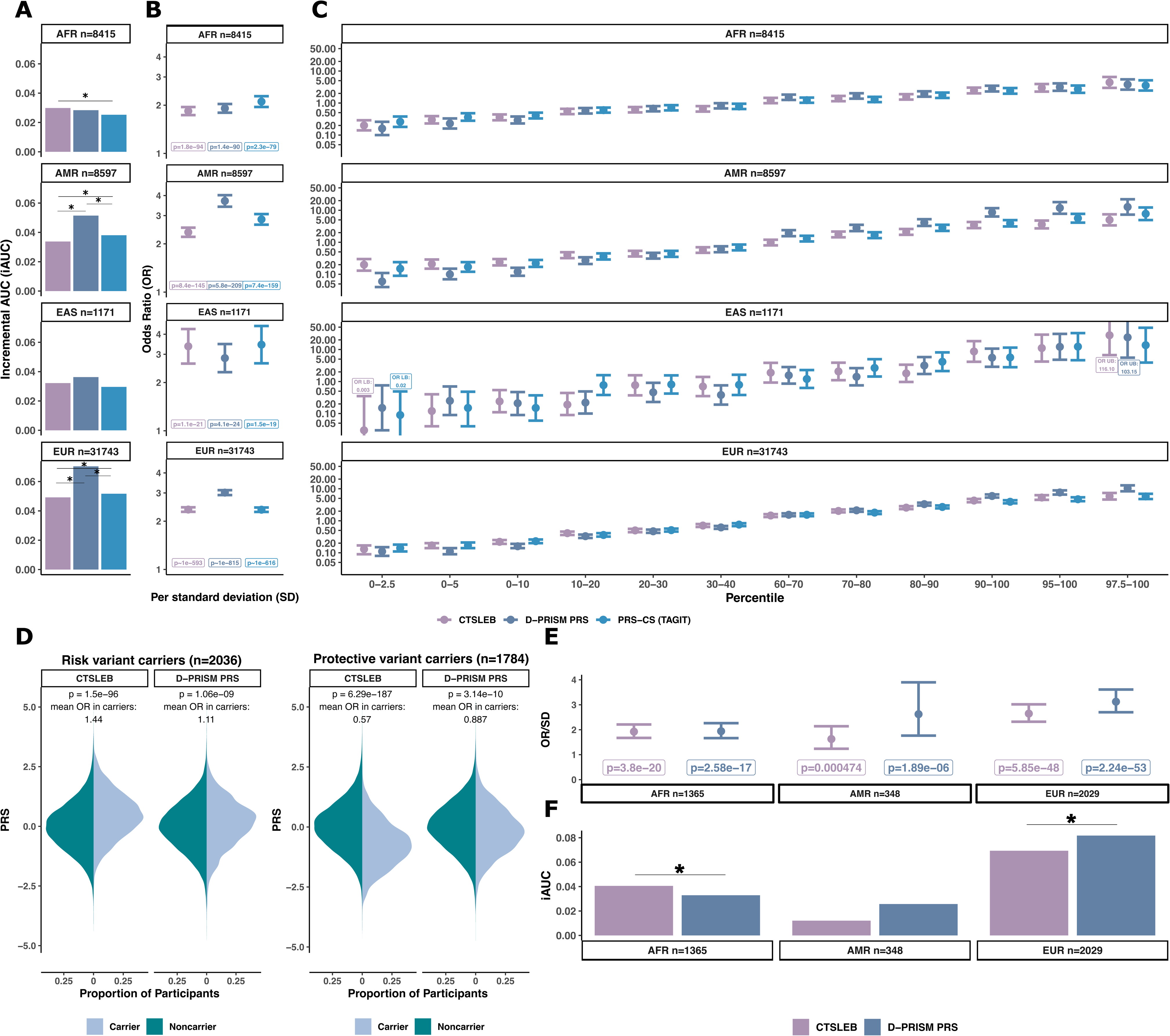
Performance of CTSLEB against “best performing” PRS utilizing larger meta-analysis (D-PRISM PRS) in validation cohort. A-C) *Performance in full validation set.* A) Incremental AUC (iAUC) of T2D predictive performance for participants in each subset ancestry. Statistical significance in AUC difference tested with DeLong test, with significance (p **≤** 0.05) denoted by *. **B)** Odds ratio (OR) of T2D risk increase per standard deviation (SD) of PRS for participants in each subset ancestry. Association p-values written below each OR/SD estimate. **C)** OR comparing PRS percentile bins to 40th-60th percentile reference group for AFR, AMR, EAS, and EUR-subset populations. **D)** Mirrored PRS distributions for carriers versus non-carriers of risk and protective variants, respectively, for CTSLEB and the D-PRISM PRS. Significance in distribution differences between carriers and non-carriers tested by the Wilcoxon rank sum, with P-values written above each mirrored distribution. The mean estimated OR associated with being a carrier versus non carrier is written above each mirrored distribution. Colors represent carrier status: dark teal for noncarriers, light blue for carriers. E-F) *Performance in rare variant carriers.* E) Odds ratio (OR) of T2D risk increase per standard deviation (SD) of PRS for rare variant carriers for AFR, AMR, EUR sub-populations. **F)** Incremental AUC (iAUC) of T2D predictive performance for rare variant carriers in subset ancestries (AFR, AMR, EUR). Statistical significance in AUC difference tested with DeLong test, with significance (p **≤** 0.05) denoted by *. Colors for A-C, D-E represent PRS used: purple for CTSLEB, heather blue for D-PRISM PRSs, bright blue for PRS-CS (TAGIT). Error bars show 95% confidence intervals. Number of participants in each subset given above each plot in A-C and below each plot for E and F.

#### D-PRISM PRS yielded higher predictive performance than PRSs with expanded variant capture

We first compared CTSLEB and the D-PRISM PRS. The D-PRISM PRS yielded higher predictive performance in AMR (CTSLEB AUC=0.854, iAUC=0.033 vs D-PRISM PRS AUC=0.872, iAUC=0.051, p(deLong test)=1.96×10^−16^) and EUR (CTSLEB AUC=0.830, iAUC=0.049 vs D-PRISM PRS AUC=0.851, iAUC=0.07, p(deLong test)=4.38×10^−49^) populations. We did not observe significant differences in the predictive performance in AFR (CTSLEB AUC=0.786, iAUC=0.30 vs D-PRISM PRS AUC = 0.784, iAUC=0.028, p(deLong test)>0.05) and EAS (CTSLEB AUC=0.890, iAUC=0.032 vs D-PRISM PRS AUC = 0.894, iAUC=0.036, p(deLong test)>0.05) (Figure 5A, Supplementary Table 5).

When comparing the D-PRISM PRS and PRS-CS (TAGIT), the D-PRISM PRS also yielded higher predictive performance in AMR (D-PRISM PRS AUC=0.872, iAUC=0.051 vs PRS-CS (TAGIT) AUC=0.859, iAUC=0.038, p(deLong test)=1.79×10^−12^) and EUR (D-PRISM PRS AUC=0.851, iAUC=0.07, PRS-CS (TAGIT) AUC=0.833, iAUC=0.052, p(deLong test)=5.90×10^−57^) populations. Similarly to the comparison with CTSLEB, we did not observe significant differences in the predictive performance in AFR (D-PRISM PRS AUC = 0.784, iAUC=0.028 vs PRS-CS (TAGIT) AUC=0.781 iAUC=0.025, p(deLong test)>0.05) or EAS (D-PRISM PRS AUC = 0.894, iAUC=0.036, PRS-CS (TAGIT) AUC=0.887, iAUC=0.30, p(deLong test)>0.05) (Figure 5A, Supplementary Table 5).

One SD of the D-PRISM PRS was also the most strongly associated with T2D status in AMR, EAS, and EUR populations, compared to CTSLEB and PRS-CS (TAGIT) with higher OR per SD increase in PRS in AMR and EUR populations. In AFR, CTSLEB showed the most significant associations with T2D status (Figure 5B, Supplementary Table 5).

The highest percentile groups for D-PRISM PRS were associated with the highest OR risk for T2D in AMR, EAS, and EUR populations when compared against both CTSLEB and PRS-CS (TAGIT). However, the highest percentile group for CTSLEB was associated with the highest OR risk for T2D in the AFR and EAS populations (Figure 5C, Supplementary Table 5).

### Capturing rare variant effects improved predictive performance in AFR rare variant carriers relative to the D-PRISM PRS

We measured whether CTSLEB performed better for carriers of rare variants than the D-PRISM PRS to test our hypothesis that performance would be improved particularly for rare variant carriers with the inclusion of rare variants in PRSs. We therefore subset the population for rare variant carriers, as described above. PRS distribution for carriers versus non-carriers of risk and protective alleles was modeled for the D-PRISM PRS and CTSLEB. We found that CTSLEB modeled the effect of both risk and protective variants to a greater degree than D-PRISM PRS (Figure 5D). While D-PRISM modeled small increases and decreases in PRSs for carriers of risk and protective variants, respectively, the mean ORs for carriers remained below 1.15 and above 0.85 (Figure 5D).

Predictive performance of CTSLEB and the D-PRISM PRS was then tested in rare variant carriers. CTSLEB showed higher predictive performance in AFR rare variant carriers (CTSLEB AUC=0.775, iAUC = 0.041 vs D-PRISM PRS AUC=0.768, iAUC=0.032, p(deLong test)=8×10^−3^) and was more significantly associated with T2D status [CTSLEB OR/SD =1.924 (1.779-2.146), p=3.8×10^−20^]. D-PRISM PRS had higher predictive performance than CTSLEB in the EUR rare variant carrier population (CTSLEB AUC=0.833, iAUC=0.069 vs D-PRISM PRS AUC=0.845, iAUC=0.081, p(deLong test)=0.042). There was no significant difference in predictive performance for the AMR rare variant carrier population (CTSLEB AUC=0.839, iAUC=0.012, D-PRISM PRS AUC=0.853, iAUC=0.026, p(deLong test)>0.05), and the D-PRISM PRS had stronger association with T2D status for both the AMR and EUR rare variant carrier populations (Figure 5E,F). PRS distributions for individual rare variant carriers were also modeled, subset by ancestry (Supplementary Figures 2-4, Supplementary Table 7). Predictive performance for samples enriched with each of these carriers was also tested, as described above. CTSLEB had higher predictive performance for carriers of all individual rare variants in AFR populations, and the D-PRISM PRS had higher predictive performance for carriers of all individual rare variants in the EUR and AMR population (Supplementary Figure 2, Supplementary Table 8).

### Capturing rare variant effects with CTSLEB increased association with age of diagnosis, particularly in rare variant carriers

To evaluate whether PRSs incorporating expanded variant frequencies and reference panels are associated with stronger genetic liability to T2D, we tested the CTSLEB-derived PRS, PRS-CS models and the benchmark D-PRISM PRSs against age at T2D diagnosis among cases. For the full population, CTSLEB was more significantly associated with decreased age of diagnosis than PRS-CS comparisons (using HapMap3 and TAGIT SNP lists) (CTSLEB p=1.9×10^−14^, PRS-CS (HM3) p=5.4×10^−10^, PRS-CS (TAGIT) p=3.3×10^−11^, respectively) and less significantly associated with decreased age of diagnosis than the benchmark D-PRISM comparison for AMR, and EUR (Figure 6A), with marginal differences between CTSLEB and D-PRISM PRS in AFR. Participants in the highest percentile bins of CTSLEB have a slightly lower age of onset in general (Figure 6B). When the population was subset to only include rare variant carriers, CTSLEB was the only PRS significantly associated with decreased age of diagnosis (p=3.0×10^−4^, beta = −0.60), with a significant association with age of diagnosis in AFR participants but not EUR participants (Figure 6C). Similarly to the full population, participants in the highest CTSLEB percentile groups had lower age of T2D onset (Figure 6D).

**Fig. 6.**
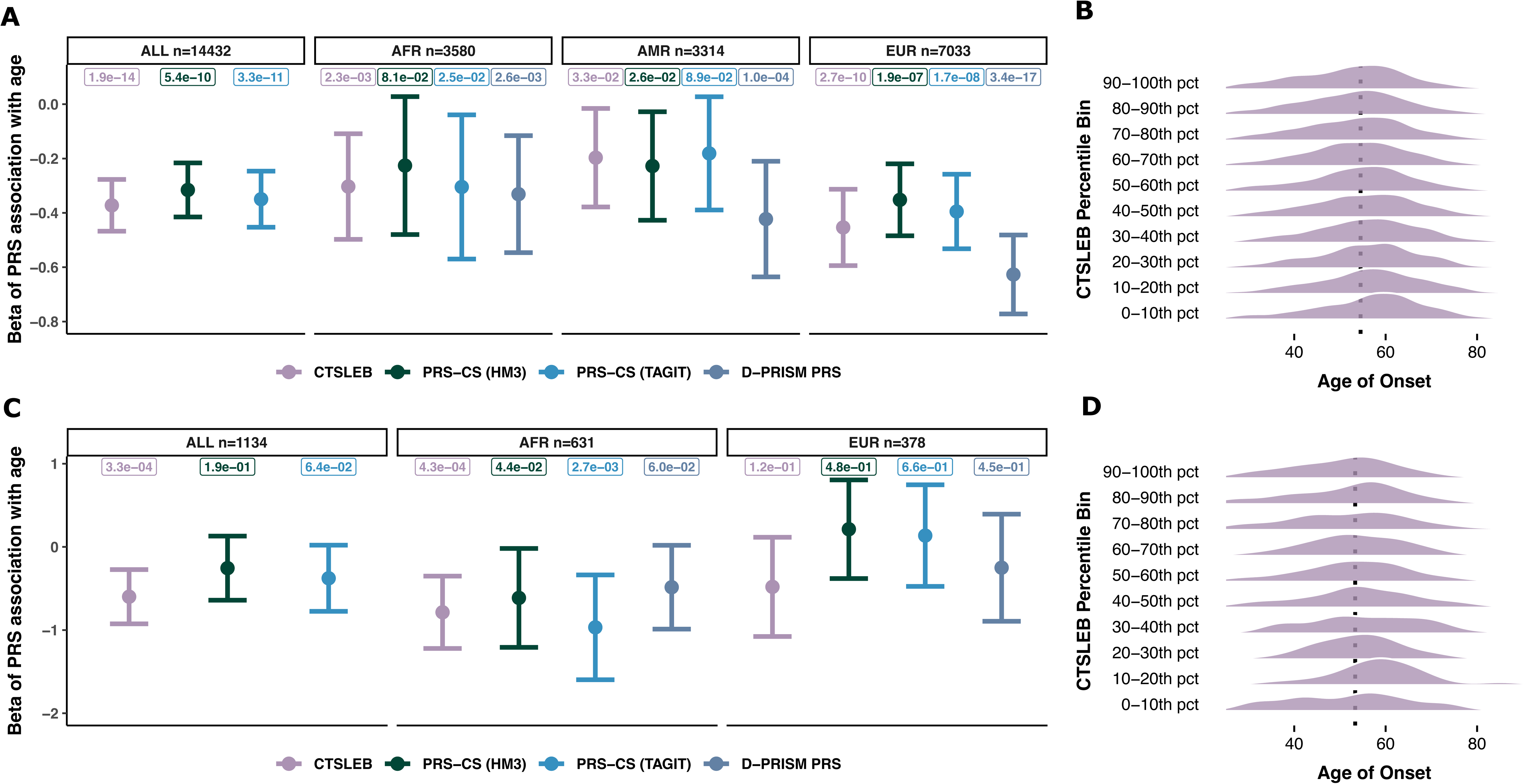
Association between PRS and age of T2D diagnosis for CTSLEB and all comparisons. A-B) *Performance in all validation set participants.* A) Beta per standard deviation (SD) association between each PRS and age of diagnosis subset in ALL, AFR, AMR, and EUR participants. Number of participants with age of T2D diagnosis shown with each ancestry. D-PRISM PRS association removed from ALL participants, as D-PRISM PRSs are subset by ancestry. 95% confidence intervals and p-value of association are shown. B) Age of diagnosis distributions for each percentile group in CTSLEB. Average age of T2D onset for the 40-60th percentile group is denoted as the dotted line. C-D) *Performance in rare variant carriers* C) Beta per standard deviation (SD) association between each PRS and age of diagnosis subset in ALL, AFR, and EUR rare variant carriers. Number of rare variant carriers with age of T2D diagnosis shown with each ancestry. D-PRISM PR association removed from ALL carriers, as D-PRISM PRSs are subset by ancestry. 95% confidence intervals and p-value of association are shown. D) Age of diagnosis distributions for each percentile group in CTSLEB in rare variant carriers. Average age of T2D onset for the 40-60th percentile group of rare variant carriers is denoted as the dotted line.

## Discussion

Identification of individuals at high risk for T2D can give critical information to patients and clinicians to help prevent or delay disease onset^41^. Polygenic risk scores, in addition to clinical risk factors, help to identify those who may develop T2D at an early age or with few clinical risk factors^42,43^. In this study, we show that expansion of variant weight sets to include rare, low-frequency and population-specific variants either with PRS-CS with an expanded TAGIT SNP list or with an adaptation of the CTSLEB framework, improves prediction accuracy. A crucial factor for these methods to work, is the development of summary statistics from large-scale GWAS meta-analysis incorporating accurate estimates from rare variants using WGS and accurate rare variant imputation.

CTSLEB showed significantly higher predictive performance for AFR participants. We think that the differences in predictive performance could be attributed to the ability to capture population-specific variants, such as chr19:41341832:G:A in *TGFB1*. This variant has an allele frequency of 0.4% in the full AoU dataset, but an AFR-specific allele frequency of 8.1%. As such, this variant is not tagged by either the HapMap3 variant list or TAGIT variant list. With an OR estimated by our meta-analysis to be 1.36, carriers of this variant were estimated to have an average OR based on the PRS value of 1.58 by CTSLEB. In contrast, PRS-CS (HM3) and PRS-CS (TAGIT) substantially underestimated risk, with mean estimated ORs based on the PRS of 1.08 and 1.02, respectively. As a result, sample populations enriched with carriers of this variant had better prediction of T2D by CTSLEB than PRS-CS methods. This highlights a critical path for achieving robust multi-ancestry prediction: increasing sample size is necessary, but optimizing methodology to capture population-specific architecture is equally vital.

While there are many GWASs identifying variants associated with changes in T2D risk, these GWASs are overrepresented by European populations^44^. Overrepresentation by European populations can lead to PRSs that are less accurate for non-European populations, exacerbating existing public health disparities^45^. When PRSs exclude population-specific variants by imposing a MAF minimum of 1%, the effects of these variants are missed completely, as demonstrated with the variant in *TGFB1*. While the TAGIT SNPset was designed to increase tagging among variants with MAF>0.01 across major ancestries, CTSLEB required the creation of a novel reference panel comprising 79.5 million and 83,818 participants to allow modeling of LD between low-frequency and rare variants. Therefore, CTSLEB allowed the incorporation of rare variant effects in the PRS and substantially improved predictive accuracy among rare variant carriers.

Beyond improving population-level prediction accuracy, our results validate the hypothesis that PRSs can model the “monogenic-like” risk conferred by rare variants. Carriers of variants with intermediate penetrance often fall into a gap between monogenic screening and common-variant PRS. The effects of these variants are not strong enough to be classified as pathogenic for monogenic diabetes but can confer substantial increased risk^13^. This risk is missed with common variant PRSs. One such variant was a previously reported coding variant in HNF4A (chr20:44413714:C:T, p.Arg114Trp)^13^, which had an odds ratio of 7.9 in our meta-analysis. The estimated average T2D OR based on the CTSLEB PRS was ~3.10 for carriers of this variant, whereas, PRS-CS (HM3) and PRS-CS (TAGIT) substantially underestimated risk, with mean estimated ORs of 0.812 and 0.853, respectively. The minimum estimated OR based on the CTSLEB PRS for a carrier of this rare variant was 1.24. For this participant, PRS-CS (HM3) and PRS-CS (TAGIT) estimated OR risks of 0.663 and 0.70, respectively. The maximum estimated OR based on the CTSLEB PRS for a carrier of the *HNF4A* variant was 7.97. PRS-CS (HM3) and PRS-CS (TAGIT) estimated OR risks of 1.12 and 1.48, respectively, for this participant. The results of CTSLEB recapitulate the results seen in Huerta-Chagoya *et al*., 2025^18^, where carriers have different risk depending on the common variant PRS, with those in the highest tertile of common variant PRS status had ORs close to that of carriers of pathogenic MODY variants in the same gene^13^. This observed improvement is expected to apply only to summary statistics generated from WGS data or from high-quality imputation, such as with the TOPMed reference panel, which we have shown provides high alternate-allele concordance for variants with MAF greater than 1×10^−4^ ^13^.

While these rare variants could be modeled separately and combined with a common variant PRS, it is advantageous to incorporate rare variants into a PRSs rather than estimating the effects of rare variants separately and linearly combining the risk, as rare variant effects are not always known and cannot always all be taken into account easily, for example, for people that carry more than one rare variant with large effect or carry multiple interacting rare variants.

To further evaluate the performance of CTSLEB in carriers of rare variants, we selected a set of low-frequency or rare variants, with modest effect sizes (absolute log-odds ratio > 0.3) and weighted average MAF < 0.03 in our meta-analysis. Carriers of our selected rare variants comprised a substantial portion of the validation cohort: almost 7.5%. Since these variants are not tagged by the HM3 or TagIt variant list, genetic risk for T2D for these participants would be underestimated by the current most widely used PRSs. CTSLEB did model the effects of these rare variants more accurately compared to other methods. CTSLEB assigned significantly higher and lower scores for carriers of risk and protective rare variants, respectively, when compared to non-carriers, whereas PRS-CS comparisons showed much smaller changes in estimated risk between carriers and non-carriers. The ability to capture these changes in risk increased predictive performance for CTSLEB, with significantly higher AUCs for CTSLEB than either PRS-CS (TAGIT) or PRS-CS (HM3) in rare variant carriers.

While the comparison of several PRS approaches methods using the same summary statistics showed promising results for PRS-CS (TAGIT) and CTSLEB, we compared then these results with the recently available PRSs developed by the D-PRISM consortium, based on common variants, but using a multi-ancestry approach and summary statistics with effective sample size 1.6-fold larger than our rare variant meta-analysis. When we compared PRS-CS (TAGIT) and CTSLEB against the D-PRISM PRS, we found the D-PRISM PRS still had the highest prediction performance and association with T2D in every ancestry except AFR. We expect that most of the differences in predictive performance can be attributed to differences in sample sizes used to construct meta-analysis summary statistics, which results in more accurate effect sizes for common variants, improving PRS performance. However, the fact that in AFR, CTSLEB showed indistinguishable performance, suggests that the improvement in variant coverage by CTSLEB may be able to overcome the larger sample size on which the D-PRISM PRS is based. CTSLEB still modeled changes in risk associated with carrying rare variants more accurately than the D-PRISM PRS and in AFR rare variant carriers, and CTSLEB showed stronger association with T2D and higher predictive performance.

Previous research has demonstrated that rare, large-effect variants strongly influence the extreme ends of complex-trait distributions, and could have an impact on the extreme liability of T2D or in cases of more severe T2D^46^. We estimated age at onset of T2D as a proxy of T2D liability and tested this trait for association with the developed PRSs. We observed that the expansion of variants included in the PRS weights to include more low-frequency and rare-variants with the CTSLEB method improved association with T2D age of diagnosis, indicating a possible stronger association with higher genetic liability predisposition. While CTSLEB did not predict age at onset better than D-PRISM, performance was indistinguishable. CTSLEB was the only PRS with a significant association with age of T2D diagnosis in rare variant carriers. These results also suggest that the incorporation of rare variants into PRSs for quantitative traits represent a promising approach. We expect that incorporation of rare variants could better capture the extremes of such traits.

PRS-CS (TAGIT) and CTSLEB generated a single set of variant weights, easily distributed and implemented. In this regard, CTSLEB and PRS-CS (TAGIT) offer an advantage over even the benchmark D-PRISM generated PRSs^19^, which require calculating ancestry-specific PRSs and combining them appropriately in the target population, and required previously fitting each participant into a categorized ancestry group. The PRSs developed here do not require fitting participants into predefined ancestry groups, making the implementation more straightforward for participants of any population group and particularly for admixed populations. We also observe that even with a much smaller AFR sample size used for summary statistics, CTSLEB still had a higher association with T2D than the D-PRISM PRSs in the AFR population, suggesting that CTSLEB may be the optimal PRS for Africans/African Americans, as this PRS would also offer improved modeling in carriers of rare variants.

CTSLEB offers advantages over other attempts to incorporate rare variant effects into PRSs, as well^16–18^. RICE creates rare variant PRSs based on burden scores of rare variant sets and integrates these with common variant PRSs via ensemble learning^17^. Gene burden tests to incorporate rare variants into PRSs have also been applied to prediction of BMI, showing limited improvement over common variant PRSs^18^. While these approaches have been effective in quantitative traits, it relies on rare variants being identified as a part of a set in order to incorporate their effects and did not perform as well in T2D. Breast and prostate cancers PRSs have included rare variants by clustering genes where these variants may be found and linearly combining the rare variant effects and common-variant PRS^16^. The functional annotation of rare variants is necessary to capture these effects, and functional annotations for rare variants are not always immediately available. The implementation of these existing methods requires pre-existing knowledge of rare variant effects and is less straightforward to implement than either CTSLEB or PRS-CS (TAGIT), particularly for non-coding variants.

This work faces some limitations, largely related to lack of diversity in cohorts used for the meta-analysis and training of PRS weights. Our meta-analysis was overrepresented by European participants, with 91% EUR participants and less than 1% MID participants. CTSLEB weights were also trained on a Latino-based cohort and applied to a multi-ancestry validation cohort. Even with a lack of diversity in the meta-analysis and training cohort, CTSLEB showed higher performance than the other approaches in the AFR population. These weights could possibly be improved with a more diverse training set and additional cohorts in the meta-analysis to increase sample sizes in underrepresented populations.

Given high predictive performance of the D-PRISM PRS utilizing large meta-analysis sample sizes and the improvement we observe through expansion of low-frequency, rare, and population-specific variants included with PRS-CS (TAGIT) and CTSLEB, we anticipate that the new wave of GWAS meta-analysis, currently underway, based on dense imputation reference panels (i.e. TOPMed) and/or WGS, will greatly benefit from the incorporation of more rare variants in PRSs. Future research should be done in the expansion of LD reference panels used with PRS-CS(x) methods to include more low-frequency variants, potentially with the creation of trait-specific LD panels that include low frequency variants known to be associated with traits of interest. These LD panels would likely still need to exclude ultra-rare variants such as the high effect p.Arg114Trp variant in *HNF4A*, but they could capture the effects of population-specific variants such as the variant in *TGFB1*. We anticipate that the approaches proposed here will continue to improve performance as larger and better meta-analyses including rare variants are generated. We believe that these methods are easily applicable to other complex diseases and quantitative traits.

In summary, this study underscores the value in incorporation of low-frequency, rare, and population-specific variants into polygenic risk score variant weight sets with high quality imputation and expanded reference panels, improving prediction performance, particularly in participants of African American ancestry and within rare variants carriers.

## Contributors

J.M.M. and K.T. designed the study. K.T. implemented the analysis and modified the T2D phenotype for AoU v8. K.T and J.M.M wrote the first draft of the manuscript. X.W. wrote the original scripts for CTSLEB clumping and training. A.H-C and M.V conducted the meta-analysis. A.H-C provided the weights for benchmark PRS comparison and created the age of T2D diagnosis phenotype. T.G and H.Z. provided advisory insight throughout analysis and study conceptualization. J.M.M directed this work. The rest of the authors provided data for different stages of the analyses. All authors contributed to interpreting the data, and they read, revised and approved the final manuscript.

## Data availability

The developed PRS weights will be available without restrictions through the PGS catalog (https://www.pgscatalog.org).

## Data Availability

Individual participant data is not available because they are subject to data protection laws and restrictions imposed by the ethics committee to ensure study participants' privacy. The study protocol and the individual methods are included in the methods section. The developed PRS weights are available without restrictions through the PGS catalog (https://www.pgscatalog.org).

## Acknowledgements

J.M.M. is supported by American Diabetes Association grant #11-22-ICTSPM-16 and by NHGRI U01HG011723, by the National Institute Of Diabetes And Digestive And Kidney Diseases of the National Institutes of Health under Award Number R01DK137993, R01DK140545 and U01 DK140757, and a Medical University of Bialystok (MUB) grant from the Ministry of Science and Higher Education (Poland). This work is supported by the Novo Nordisk Foundation (NNF21SA0072102). T.H.H is supported by the Agency for Science, Technology and Research (A*STAR, Singapore) National Science Scholarship. A.H.-C. is supported by the American Diabetes Association grant 11-23-PDF-35. This research was supported in part by the Intramural Research Program of the National Institutes of Health (NIH). The contributions of the NIH authors were made as part of their official duties as NIH federal employees, are in compliance with agency policy requirements, and are considered Works of the United States Government. However, the findings and conclusions presented in this paper are those of the authors and do not necessarily reflect the views of the NIH or the U.S. Department of Health and Human Services. This research was funded in part by Wellcome (grant no. 220540/Z/20/A, “Wellcome Sanger Institute Quinquennial Review 2021–2026”). For the purpose of open access, the authors have applied a CC-BY public copyright license to any author accepted manuscript version arising from this submission.

<u>AoU Research Program (AOU)</u> is supported by the National Institutes of Health, Office of the Director: Regional Medical Centers: 1 OT2 OD026549; 1 OT2 OD026554; 1 OT2 OD026557; 1 OT2 OD026556; 1 OT2 OD026550; 1 OT2 OD 026552; 1 OT2 OD026553; 1 OT2 OD026548; 1 OT2 OD026551; 1 OT2 OD026555; IAA #: AOD 16037; Federally Qualified Health Centers: HHSN 263201600085U; Data and Research Center: 5 U2C OD023196; Biobank:1 U24 OD023121; The Participant Center: U24 OD023176; Participant Technology Systems Center: 1 U24 OD023163; Communications and Engagement: 3 OT2 OD023205; 3 OT2 OD023206; and Community Partners: 1 OT2 OD025277; 3 OT2 OD025315; 1 OT2 OD025337; 1 OT2 OD025276. In addition, the AoU Research Program would not be possible without the partnership of its participants.

<u>Estonian Biobank (ESTBB)</u> was funded by the Estonian Research Council Grant PRG1291. All computational analyses were carried out in the High-Performance Computing Center, University of Tartu. We also acknowledge the participants of the Estonian Biobank for their contributions.

<u>FinnGen study (FINNGEN)</u> is a large-scale genomics initiative that has analyzed over 500,000 Finnish biobank samples and correlated genetic variation with health data to understand disease mechanisms and predispositions. The project is a collaboration between research organisations and biobanks within Finland and international industry partners. We want to acknowledge the participants and investigators of the FinnGen study.

<u>Resource for Genetic Epidemiology on Adult Health and Aging (GERA)</u> was supported by a grant (RC2 AG033067; PIs Schaefer and Risch) awarded to the Kaiser Permanente Research Program on Genes, Environment, and Health (RPGEH) and the UCSF Institute for Human Genetics. The RPGEH was supported by grants from the Robert Wood Johnson Foundation, the Wayne and Gladys Valley Foundation, the Ellison Medical Foundation, Kaiser Permanente Northern California, and the Kaiser Permanente National and Northern California Community Benefit Programs.

<u>Genes and Health.</u> Genes & Health is/has recently been core-funded by Wellcome (WT102627, WT210561), the Medical Research Council (UK) (M009017, MR/X009777/1, MR/X009920/1), Higher Education Funding Council for England Catalyst, Barts Charity (845/1796), Health Data Research UK (for London substantive site), and research delivery support from the NHS National Institute for Health Research Clinical Research Network (North Thames). Genes & Health is/has recently been funded by Alnylam Pharmaceuticals, Genomics PLC; and a Life Sciences Industry Consortium of AstraZeneca PLC, Bristol-Myers Squibb Company, GlaxoSmithKline Research and Development Limited, Maze Therapeutics Inc, Merck Sharp & Dohme LLC, Novo Nordisk A/S, Pfizer Inc, Takeda Development Centre Americas Inc. We thank Social Action for Health, Centre of The Cell, members of our Community Advisory Group, and staff who have recruited and collected data from volunteers. We thank the NIHR National Biosample Centre (UK Biocentre), the Social Genetic & Developmental Psychiatry Centre (King’s College London), Wellcome Sanger Institute, and Broad Institute for sample processing, genotyping, sequencing and variant annotation. This work uses data provided by patients and collected by the NHS as part of their care and support. This research utilised Queen Mary University of London’s Apocrita HPC facility, supported by QMUL Research-IT, http://doi.org/10.5281/zenodo.438045. We thank: Barts Health NHS Trust, NHS Clinical Commissioning Groups (City and Hackney, Waltham Forest, Tower Hamlets, Newham, Redbridge, Havering, Barking and Dagenham), East London NHS Foundation Trust, Bradford Teaching Hospitals NHS Foundation Trust, Public Health England (especially David Wyllie), Discovery Data Service/Endeavour Health Charitable Trust (especially David Stables), Voror Health Technologies Ltd (especially Sophie Don), NHS England (for what was NHS Digital) - for GDPR-compliant data sharing backed by individual written informed consent. Most of all we thank all of the volunteers participating in Genes & Health.

<u>Mexican Biobank (MXBB)</u> was supported by Mexico’s CONACYT (Grant number FONCICYT/50/2016; PI Moreno-Estrada), and the Newton Fund through the UK Medical Research Council (Grant number MR/N028937/1; PI Moreno-Estrada) to genetically characterize the population-based cohort derived from the National Health Survey 2000 (ENSA2000). The resulting ENSA Genomics Consortium acknowledges the seminal effort of Dr. Jaime Sepúlveda, the Mexican Ministry of Health, and the National Institute of Public Health, in the design and implementation of the ENSA2000 survey from which genomic data were generated for the MXB Project. Members of the ENSA Genomics Consortium are also acknowledged for biobank maintenance, sample selection and processing of materials contributed to the MXBB.

Mass General Brigham Biobank (MGBB) acknowledges the Partners HealthCare System for support of the MGB biobank and MGB patients for providing samples, genomic data, and health information data, as well as research support by NIDDK K24 DK110550 (to J.C.F.), K24 DK080140 (to J.B.M.) and NIDDK K23DK114551 (to M.S.U).

<u>MyCode (Geisinger)</u> recruitment and exome sequencing were funded through a partnership between Geisinger and the Regeneron Genetics Center. We thank the participants and providers for their contributions to this study.

<u>Slim Initiative for Genomic Medicine in the Americas (SIGMA).</u> This work was conducted as part of the Slim Initiative for Genomic Medicine, a joint U.S.-Mexico project funded by the Carlos Slim Health Institute. The UNAM/INCMNSZ diabetes study was supported by Consejo Nacional de Ciencia y Tecnología grants 138826, 128877, CONACyT-SALUD 2009-01-115250, and a grant from Dirección General de Asuntos del Personal Académico, UNAM, IT 214711. The Diabetes in Mexico Study was supported by Consejo Nacional de Ciencia y Tecnología grant 86867 and by Instituto Carlos Slim de la Salud, A.C. The Mexico City Diabetes Study was supported by National Institutes of Health (NIH) grant R01HL24799 and by the Consejo Nacional de Ciencia y Tenologia grants: 2092, M9303, F677-M9407, 251M, and 2005-C01-14502, SALUD 2010-2-151165. The Multiethnic Cohort was supported by NIH grants CA164973, CA054281, and CA063464.

<u>UK Biobank (UKBB)</u> analyses were conducted using the UK Biobank resource under applications 236, 9161, and 10035. This research was supported by the British Heart Foundation (grant SP/13/2/30111). Large-scale comprehensive genotyping of UK Biobank for cardiometabolic traits and diseases: UK CardioMetabolic Consortium (UKCMC).

Wellcome Trust Case Control Consortium (WTCCC) analysis and genotyping was supported by: Wellcome Trust funding 090367, 098381, 090532, 083948, 085475, 101630, and 203141; MRC (G0601261); EU (Framework 7) HEALTH-F4-2007-201413; and NIDDK DK098032 and U01-DK105535.

