## Supplementary Information for "Improving type 2 diabetes polygenic risk scores by incorporating rare, low-frequency, and population-specific variants"

**Supplementary Material**

| **Supplementary Figures** | **1** |
| --- | --- |
| **Cohort acknowledgements and funding** | **5** |
| **Genes and Health Research Team Authorship for Scientific Publications** | **7** |

**Supplementary Figures**

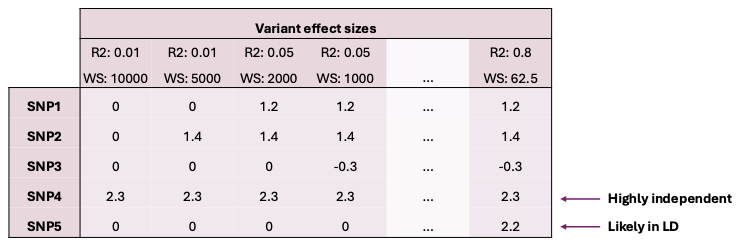

**Supplementary Fig. 1 |** Mock table of effect sizes for each LD clumping parameter. When a variant is determined to be in LD with another variant for a given LD clumping parameter, it is given a weight of 0 and therefore would not contribute to the calculation of PRS for the given variant weight-set specific PRS.

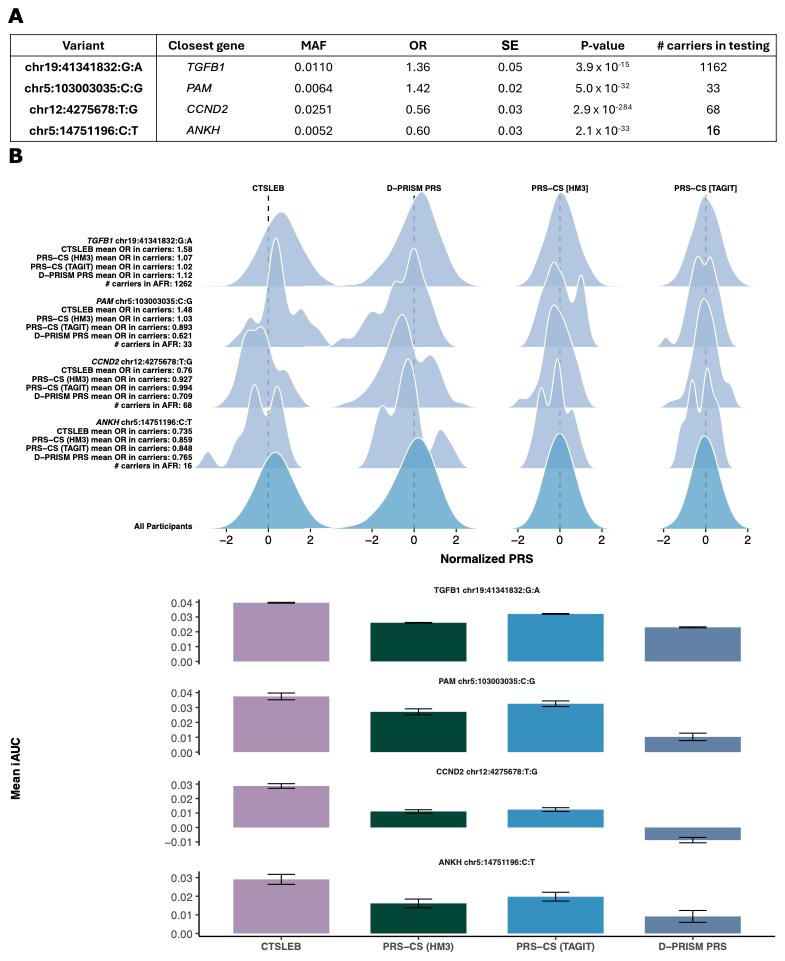

**Supplementary Fig 2 | PRS distribution for carriers of individual rare variants and predictive performance for samples enriched with carriers of rare variants in AFR.** A) Variants and associated MAFs, ORs, SEs, p-values, found by meta-analysis. Number of carriers in validation cohort. B) Ridgeline plot showing PRS distributions for carriers of rare variants for CTSLEB and PRS-CS comparisons created with the same meta-analysis. Variant, number of carriers in validation cohort, and OR being a variant carrier, as determined by the meta-analysis, are written to the left of each distribution set. Non-carrier distributions are at the bottom of the plot. C) Mean incremental AUC (iAUC) with 95% confidence interval for samples enriched with carriers. For variants with < 30 carriers in validation cohort, sample size is 4 x number of carriers. Otherwise, sample size is 2x number of carriers. Non-carriers are randomly selected and predictive performance for each sample set is tested over 100 iterations for each variant. Colors represent PRS used: purple for CTSLEB, green for PRS-CS (HM3), bright blue for PRS-CS (TAGIT), muted blue for D-PRISM PRS.

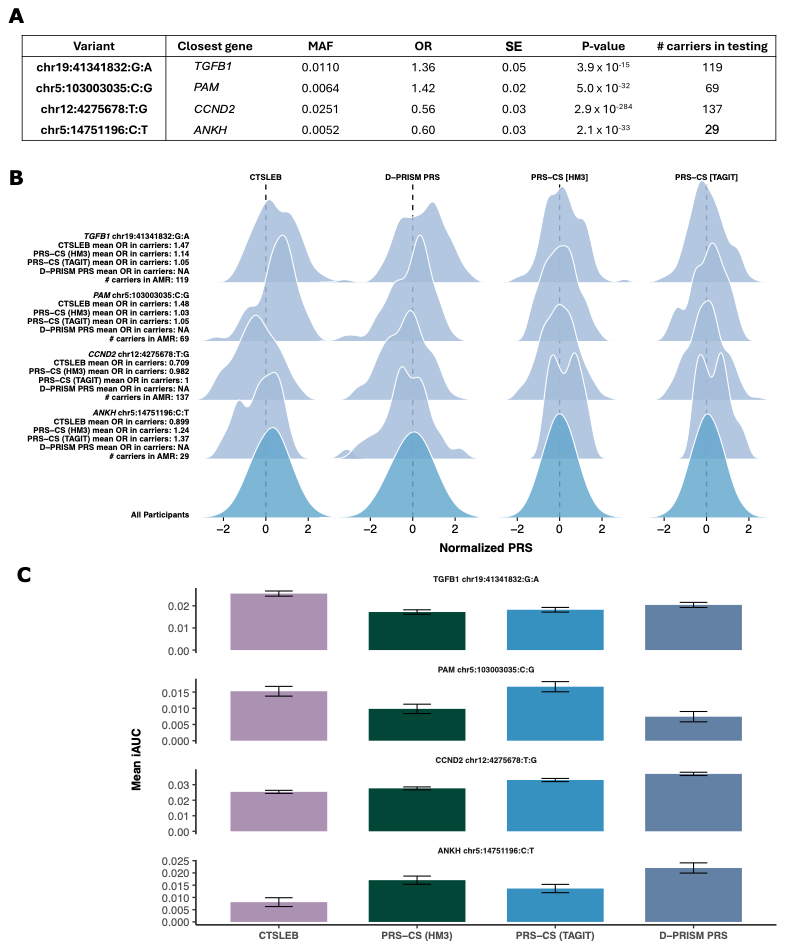

**Supplementary Fig 3 | PRS distribution for carriers of individual rare variants and predictive performance for samples enriched with carriers of rare variants in AMR.** A) Variants and associated MAFs, ORs, SEs, p-values, found by meta-analysis. Number of carriers in validation cohort. B) Ridgeline plot showing PRS distributions for carriers of rare variants for CTSLEB and PRS-CS comparisons created with the same meta-analysis. Variant, number of carriers in validation cohort, and OR being a variant carrier, as determined by the meta-analysis, are written to the left of each distribution set. Non-carrier distributions are at the bottom of the plot. C) Mean incremental AUC (iAUC) with 95% confidence interval for samples enriched with carriers. For variants with < 30 carriers in validation cohort, sample size is 4 x number of carriers. Otherwise, sample size is 2x number of carriers. Non-carriers are randomly selected and predictive performance for each sample set is tested over 100 iterations for each variant. Colors represent PRS used: purple for CTSLEB, green for PRS-CS (HM3), bright blue for PRS-CS (TAGIT), muted blue for D-PRISM PRS.

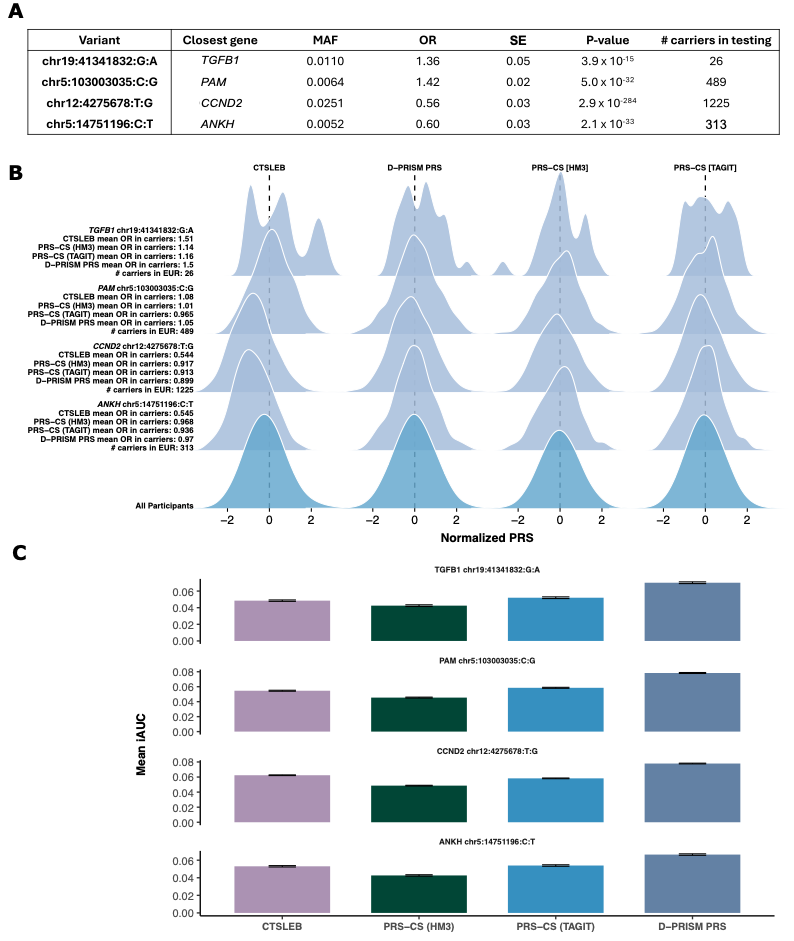

**Supplementary Fig. 4 | PRS distribution for carriers of individual rare variants and predictive performance for samples enriched with carriers of rare variants in EUR.** A) Variants and associated MAFs, ORs, SEs, p-values, found by meta-analysis. Number of carriers in validation cohort. B) Ridgeline plot showing PRS distributions for carriers of rare variants for CTSLEB and PRS-CS comparisons created with the same meta-analysis. Variant, number of carriers in validation cohort, and OR being a variant carrier, as determined by the meta-analysis, are written to the left of each distribution set. Non-carrier distributions are at the bottom of the plot. C) Mean incremental AUC (iAUC) with 95% confidence interval for samples enriched with carriers. For variants with < 30 carriers in validation cohort, sample size is 4 x number of carriers. Otherwise, sample size is 2x number of carriers. Non-carriers are randomly selected and predictive performance for each sample set is tested over 100 iterations for each variant. Colors represent PRS used: purple for CTSLEB, green for PRS-CS (HM3), bright blue for PRS-CS (TAGIT), muted blue for D-PRISM PRS.

**Cohort acknowledgements and Funding**

All Of Us Research Program (AOU) is supported by the National Institutes of Health, Office of the Director: Regional Medical Centers: 1 OT2 OD026549; 1 OT2 OD026554; 1 OT2 OD026557; 1 OT2 OD026556; 1 OT2 OD026550; 1 OT2 OD 026552; 1 OT2 OD026553; 1 OT2 OD026548; 1 OT2 OD026551; 1 OT2 OD026555; IAA #: AOD 16037; Federally Qualified Health Centers: HHSN 263201600085U; Data and Research Center: 5 U2C OD023196; Biobank:1 U24 OD023121; The Participant Center: U24 OD023176; Participant Technology Systems Center: 1 U24 OD023163; Communications and Engagement: 3 OT2 OD023205; 3 OT2 OD023206; and Community Partners: 1 OT2 OD025277; 3 OT2 OD025315; 1 OT2 OD025337; 1 OT2 OD025276. In addition, the All of Us Research Program would not be possible without the partnership of its participants.

Estonian Biobank (ESTBB) was funded by the Estonian Research Council Grant IUT20-60, IUT24-6, PRG687, and the European Union through the European Regional Development Fund Project No. 2014-2020.4.01.15-0012 GENTRANSMED.

**Genes & Health Research Team authorship for Scientific Publications**

Eamonn Maher	Aston University

Shabana Chaudhary Blizard Institute, Queen Mary University of London

Joseph Gafton	Blizard Institute, Queen Mary University of London

Karen A Hunt	Blizard Institute, Queen Mary University of London

Shapna Hussain	Blizard Institute, Queen Mary University of London

Kamrul Islam	Blizard Institute, Queen Mary University of London

Mohammed Bodrul Mazid	Blizard Institute, Queen Mary University of London

Elizabeth Owor	Blizard Institute, Queen Mary University of London

Jessry Russell Blizard Institute, Queen Mary University of London

Nishat Safa	Blizard Institute, Queen Mary University of London

John Solly	Blizard Institute, Queen Mary University of London

Marie Spreckley Blizard Institute, Queen Mary University of London

David A Van Heel	Blizard Institute, Queen Mary University of London

Jan Whalley	Blizard Institute, Queen Mary University of London

Ishevanhu Zengeya	Blizard Institute, Queen Mary University of London

Emily Mantle	Blizard Institute, Queen Mary University of London

Shaheen Akhtar	Bradford Teaching Hospitals NHS Foundation Trust

Samina Ashraf	Bradford Teaching Hospitals NHS Foundation Trust

Dan Mason Bradford Teaching Hospitals NHS Foundation Trust

John Wright Bradford Teaching Hospitals NHS Foundation Trust

Daniel MacArthur	Garvan Institute

Michael Simpson	King's College London

Richard C Trembath	King's College London

Gerome Breen Kings College London

Raymond Chung Kings College London

Sang Hyuck Lee Kings College London

Omar Asgar	Manchester University Hospitals

Joanne Harvey Manchester University Hospitals

Karen Tricker Manchester University Hospitals

Caroline Winckley	Manchester University Hospitals

Hanifa Khatun Manchester University Hospitals

Amna Asif	Manchester University Hospitals

Claudia Langenberg Precision Healthcare University Research Institute, Queen Mary University of London

Grainne Colligan	Social Action for Health (charity)

Ceri Durham	Social Action for Health (charity)

| Bill Newman | University of Manchester |
| --- | --- |
| Ahsan Khan | Waltham Forest Council |
| Hilary Martin | Wellcome Sanger Institute |
| Teng Heng | Wellcome Sanger Institute |
| Matt Hurles | Wellcome Sanger Institute |
| Vivek Iyer | Wellcome Sanger Institute |

Georgios Kalantzis	Wellcome Sanger Institute

Vladimir Ovchinnikov	Wellcome Sanger Institute

| Iaroslav Popov Wellcome Sanger Institute  Klaudia Walter Wellcome Sanger Institute  Panos Deloukas William Harvey Research Institute, Queen Mary University of London | |
| --- | --- |
| David Collier | William Harvey Research Institute, Queen Mary Universityof London |
| Ana Angel of London | Wolfson Institute of Population Health, Queen Mary University |
| Saeed Bidi London | Wolfson Institute of Population Health, Queen Mary University of |
| Fabiola Eto | Wolfson Institute of Population Health, Queen Mary University of London |
| Sarah Finer London | Wolfson Institute of Population Health, Queen Mary University of |
| Chris Griffiths  London | Wolfson Institute of Population Health, Queen Mary University of |
| Sam Hodgson | Wolfson Institute of Population Health, Queen Mary University of |

London

Benjamin M Jacobs Wolfson Institute of Population Health, Queen Mary University of London

Rohini Mathur	Wolfson Institute of Population Health, Queen Mary University of

London

Caroline Morton Wolfson Institute of Population Health, Queen Mary University of London

Asma Qureshi Wolfson Institute of Population Health, Queen Mary University of London

Stuart Rison	Wolfson Institute of Population Health, Queen Mary University of

London

Annum Salman	Wolfson Institute of Population Health, Queen Mary University of

London

Miriam Samuel Wolfson Institute of Population Health, Queen Mary University of London

Moneeza K Siddiqui Wolfson Institute of Population Health, Queen Mary University of London

| Daniel Stow London | Wolfson Institute of Population Health, Queen Mary University of |
| --- | --- |
| Sabina Yasmin of London | Wolfson Institute of Population Health, Queen Mary University |
| Julia Zöllner  London | Wolfson Institute of Population Health, Queen Mary University of |
| Sheik Dowlut London | Wolfson Institute of Population Health, Queen Mary University of |
